# RadGuide AI: Development and Technical Evaluation of a General Nuclear Medicine Agent for Traceable Radiopharmaceutical Decision Support

**DOI:** 10.64898/2026.07.09.26357614

**Authors:** Xiaochun Gu, Haidong Zhu, Fei Zhong, Gao-Jun Teng

## Abstract

**Background:** Nuclear medicine and radiopharmaceutical development require coordinated radiochemistry, dosimetry, molecular imaging, radiation-safety and clinical decision processes. Current workflows remain fragmented, difficult to audit and poorly standardised for evaluating domain-specific AI support.

**Methods:** We developed RadGuide AI, a nuclear medicine agent built around a traceable data-model-tool loop. Patent, literature and clinical-trial records were converted into 15,596 initial QA items; relevance screening, completeness checks, semantic deduplication and cross-validation retained 5,474 core QA items. MedGemma-27B-Instruct served as the foundation model and was adapted with LoRA. The system incorporated 55 MCP-wrapped tools covering radiopharmaceutical R&D, clinical decision support, imaging analysis and radiation-safety/dosimetry. Evaluation used a locked N=200 benchmark with predefined denominators, leakage control, expert scoring, statistical procedures, factuality audits and tool-execution metrics.

**Results:** RadGuide-LLM achieved 88.5% answer accuracy (177/200; 95% CI, 83.3-92.2%) and a Macro-Average score of 21.5/25 (bootstrap 95% CI, 20.9-22.0), exceeding GPT-4o, DeepSeek-V3.2 and the base MedGemma model in this technical evaluation. Supplementary audits reported guideline compliance, terminology recall, knowledge coverage, tool-routing success and preclinical/phantom dosimetry agreement with explicit denominators and confidence intervals.

**Interpretation:** RadGuide AI converts nuclear medicine queries into auditable retrieval, tool selection, calculation, verification and reporting workflows. The findings support technical feasibility, not definitive patient-level clinical validation; prospective multicentre studies and external benchmark release remain required before clinical deployment.

## Introduction

Artificial intelligence research has long pursued systems capable of autonomous learning, reasoning, and solving complex tasks. The rapid advancement of large language models (LLMs), exemplified by ChatGPT ^1^, has catalysed a paradigm shift in natural language processing, giving rise to novel methodologies such as chain-of-thought reasoning, retrieval-augmented generation (RAG), multimodal interaction, and collaborative reasoning among agents. Against this backdrop, AI agents have progressively evolved into interactive systems capable of coordinating large language models, specialised tools, and experimental platforms. Through task decomposition, plan generation, tool invocation, and reflective error correction, they establish a closed-loop learning mechanism of “reasoning-action-verification”. The healthcare sector is regarded as a pivotal application domain for large models ^2–4^. Although domain-adapted models such as ChatDoctor ^5^, BioMistral ^6^, and Med-PaLM 2 ^7^ have emerged with promising performance in certain benchmarks, their training data and methodologies remain largely undisclosed, limiting the reliability and reproducibility of clinical applications. A more fundamental contradiction lies in general-purpose LLMs’ lack of internalised specialised mechanisms and quantitative rules. This predisposes them to produce unsubstantiated “hallucinatory” outputs, failing to meet healthcare decision-making’s stringent requirements for evidence sufficiency, numerical precision, procedural compliance, and risk control. This issue is particularly acute given the scarcity of high-quality medical data. Nuclear medicine and radiopharmaceutical development are particularly sensitive to these challenges ^8^. Their end-to-end workflow encompasses radiochemical synthesis, molecular imaging quantification, dosimetric assessment, radiation safety, and regulatory compliance—featuring highly engineered characteristics: Firstly, critical conclusions rely on physical and numerical computations (e.g., nuclear decay parameters, energy deposition, dose estimation), demanding strict unit consistency and controllable error margins; Secondly, decision-making must adhere to clinical guidelines, standard operating procedures (SOPs), and safety thresholds, emphasising process interpretability and auditability. Finally, task execution relies on the coordinated operation of complex toolchains, including image segmentation/registration, dosimetry engines, and simulation tools. Consequently, the radiopharmaceutical domain urgently requires an intelligent system capable of operating within an evidence-computational closed loop, rather than merely functioning as a text-based question-answering model.

Addressing these requirements, we developed RadGuide AI. Its design follows a data-model-tool closed-loop principle: (1) Data layer: a technology gap-driven strategy constructs training and evaluation corpora from patents, literature, and clinical-trial records with source-level hold-out and semantic leakage control; (2) Model layer: MedGemma-27B-Instruct is adapted with LoRA to improve nuclear medicine terminology, structured reasoning, and numerical answer generation; (3) Tool layer: 55 MCP tools support radiopharmaceutical development, imaging analysis, dose calculation, and safety assessment. The system records evidence sources, tool calls, parameters, verification checks, and outputs for auditability.

RadGuide AI is intended as a human-in-the-loop research and decision-support prototype. It does not replace clinical judgement; all dosing-relevant or safety-critical outputs require review by qualified clinicians or medical physicists.

The principal contributions of this paper are as follows:

1. Proposing RadGuide AI, an agent framework for the entire radiopharmaceutical chain, which employs a “data-model-tool” closed-loop to achieve evidence traceability, computation executability, and process auditability in decision support.
2. Constructs domain-specific data assets and evaluation benchmarks for nuclear medicine: Through technology gap-driven multi-source data integration and progressive quality control processes, it generates high-confidence training data and benchmarks, providing a unified evaluation standard for enhancing domain model capabilities.
3. Achieved domain-specific fine-tuning and toolchain integration: Employed LoRA fine-tuning on a general medical foundation, while integrating 55 radiopharmaceutical R&D tools via MCP. This covers R&D, imaging, dosimetry, and safety compliance modules, enabling closed-loop execution of complex tasks through “retrieval-computation-validation-generation”.

## Methods and Results

### Architectural Design

RadGuide AI is a universal intelligent agent system for radiopharmaceutical development and nuclear medicine applications. Its overall methodology employs a “data-model-tool” closed-loop and layered decoupling approach, forming a traceable, computable, and auditable end-to-end decision support architecture (see Figure 1).

**Figure 1.**
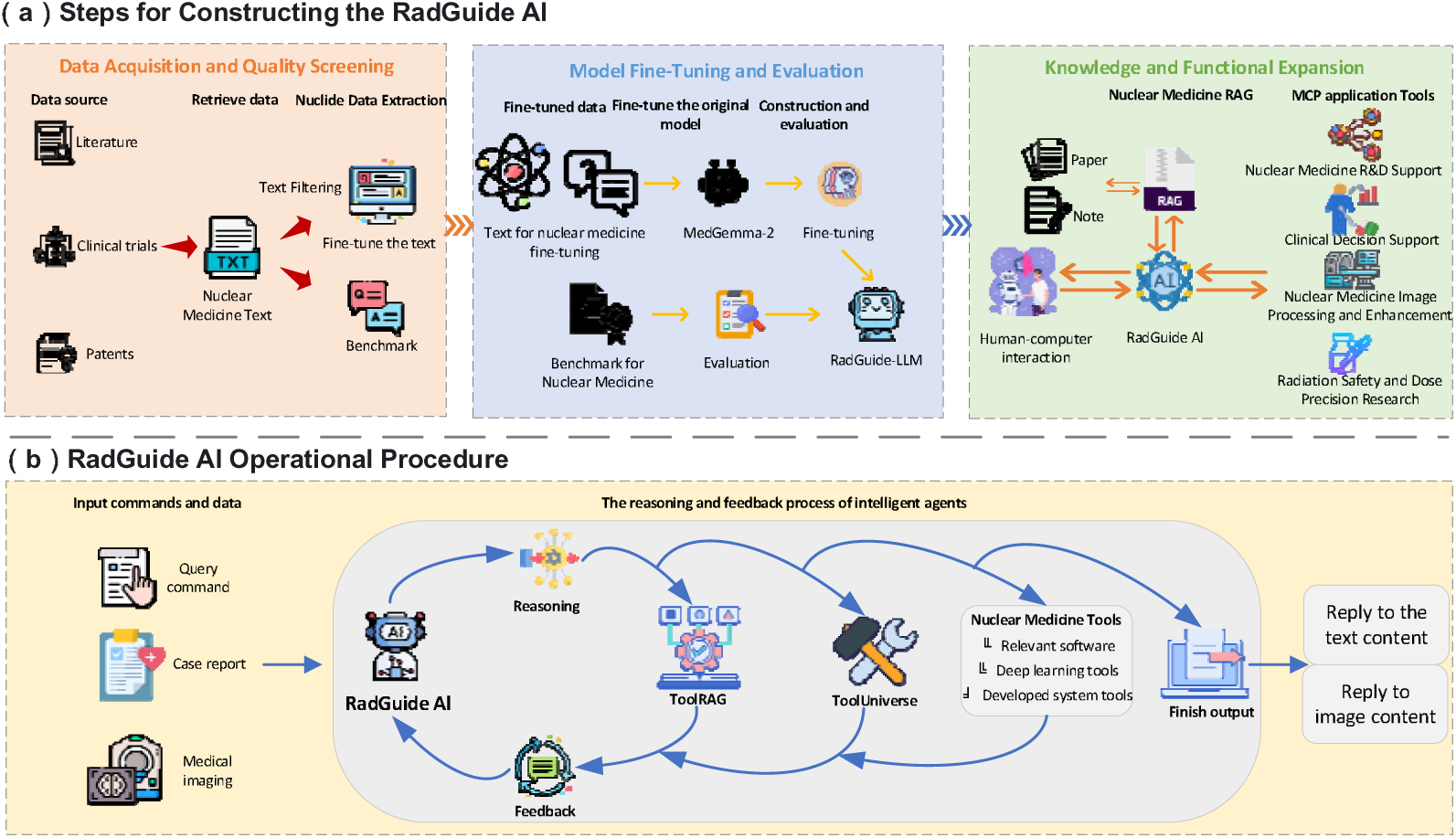
RadGuide AI system architecture and technical implementation process

The system comprises four core modules: (i) Multi-source Data & Benchmark: Addressing the full-chain capability requirements of nuclear medicine, it integrates data from three sources—patents, literature, and clinical trials—to generate task-specific datasets. Through a “technology gap-driven” data construction and quality control process, it produces highly reliable training corpora. Concurrently, it establishes domain-specific Benchmark and metrics for unified training and reproducible evaluation. (ii) Domain-Specific Cognitive Core (RadGuide-LLM): Built upon the MedGemma-27B-Instruct foundation, this employs efficient LoRA parameter fine-tuning to achieve radionuclide knowledge transfer and numerical reasoning enhancement. This endows the model with domain-consistent output capabilities for tasks including radionuclide selection, dose reasoning, and clinical question answering. (iii) Three-Tier Collaborative Agent Architecture (Agent Orchestration) ^11^: Constructs a tripartite collaborative framework comprising the Nuclear Physics Tool Universe (NucToolUniverse) — Radiopharmaceutical Large Language Model (RadGuide-LLM) — Intelligent Decision Engine (NucDecision Engine), achieving deep integration of “linguistic reasoning + specialised computation + decision orchestration“; The decision engine handles task decomposition, tool selection, and multi-step execution while maintaining auditable trails for critical steps. (iv) Medical Intelligence System and Closed-Loop Execution (RAG + Tools + Workflow): Adopting knowledge augmentation and tool-based execution as implementation pathways, this approach builds a dynamic knowledge base via RAGFlow. It integrates and schedules radiopharmaceutical R&D tools through the MCP, covering four modules: R&D support, clinical decision-making, medical imaging processing, and radiological safety/Precision Dose Calculation. This forms an executable chain of “Retrieval-Calculation-Verification-Generation”, transforming complex radiopharmaceutical tasks into reproducible workflow outputs.

At the implementation level, RadGuide AI integrates 55 MCP tools to instrument key capabilities including dosimetric simulation, image analysis, pharmacokinetic modelling, and radiation safety assessment. This enables the system to dynamically invoke external computational modules during multi-step reasoning while maintaining full-chain documentation of evidence sources and computational processes, meeting transparency and auditability requirements for high-risk scenarios.

#### RadGuide Large Language Model

RadGuide-LLM, the model component within the intelligent agent system, is built on MedGemma-27B-Instruct and adapted to nuclear medicine tasks through parameter-efficient LoRA fine-tuning. The model uses FP16/BF16 mixed-precision training to balance numerical stability and GPU memory use. LoRA rank, dropout, and target modules were selected using validation-set loss stability and benchmark performance. Claims requiring external clinical cohorts, such as cross-centre decision-error reduction or patient-level dose-bias reduction, were removed unless supported by a prespecified validation dataset.

Training used gradient accumulation to approximate an effective batch size of 16, together with warm-up, cosine learning-rate decay, and gradient clipping to stabilise optimisation. Retrieval-Augmented Generation was implemented through a three-tier knowledge base covering literature, clinical trials, and patents. Term recall, knowledge coverage, command recognition, and reasoning accuracy are reported only when denominators, scoring rubrics, and confidence intervals are specified.

#### Intelligent Decision Engine

As the central control hub of the RadGuide AI system, the Intelligent Decision Engine employs a dynamic tool selection mechanism based on the TOOLRAG model. It programmatically retrieves relevant tool sets from the nuclear physics tool universe by analysing task semantics. The engine innovatively integrates the ReAct (Reasoning+Acting) inference framework, decomposing complex clinical tasks such as “nuclear medicine R&D feasibility assessment 13,14” into a chained reasoning process: “isotope selection → labelling efficiency prediction → dose safety verification → clinical suitability analysis”. Each step follows a closed-loop logic of “think-act-observe” while fully documenting the reasoning trajectory. The system achieves real-time data synchronisation with the IAEA nuclide database via API interfaces, ensuring the timeliness of critical information such as nuclide decay parameters. Concurrently, it establishes standardised tool description specifications, clearly defining the input/output parameters and data type requirements for each tool.

The decision engine incorporates a multimodal data-fusion workflow capable of coordinating radionuclide decay curves, PET/SPECT images, molecular structure SMILES strings, and electronic health records. It supports cross-modal reasoning through feature transformation and association modelling. Safety controls include dose-calculation and threshold-verification checks, risk alerts when predefined limits are exceeded, and audit logs for any recommended adjustment. Timed workflow-efficiency claims are treated as future evaluation endpoints requiring a prespecified comparator, user cohort, task set, and statistical procedure.

#### Nuclear Medicine Tool Applications

To address the multidimensional requirements of nuclear medicine R&D, clinical decision support, imaging analysis, and radiation-safety management, we designed a modular nuclear medicine tool system. The system is centred on auditable computation, evidence retrieval, and tool coordination, and integrates physics-based algorithms, deep learning models, and domain expertise as callable modules. It provides a traceable workflow from radiopharmaceutical molecular design to personalised therapy support and radiation-safety management, with qualified clinician or medical-physicist review required for safety-critical outputs.

In supporting the entire nuclear medicine R&D process, this system implements a computation-driven research paradigm. Taking target information and therapeutic objectives as inputs, the system sequentially invokes nuclide property matching tools, labelling chemistry simulation tools, and in vitro stability assessment tools via an intelligent decision engine. This completes end-to-end simulation spanning nuclide selection, conjugation scheme optimisation, and pharmacokinetic prediction. The entire process incorporates multi-step reasoning and risk alert mechanisms. Should critical metrics (such as labelling efficiency, in vitro stability, or radiation dose) fall below preset thresholds, the system generates structured reports proposing strategies including radionuclide substitution and linker optimisation. This forms a closed-loop R&D cycle of “design-simulation-verification-optimisation”, reducing the traditional trial-and-error screening cycle.

For clinical decision support, the system focuses on personalised dose calculation and structured decision support. Based on patient clinical parameters and radiopharmaceutical properties, it can generate draft activity-regimen estimates for qualified review and organ dose distributions through the coordinated operation of automated organ segmentation, MIRD dose 16 coefficient lookup, and absorbed dose calculation engines. For special patient populations, the system incorporates physiological pharmacokinetic models and a radiation safety threshold database to enable automatic dose adjustments and contraindication alerts. During diagnostic evaluation, integrated quantitative imaging analysis tools and clinical guideline knowledge bases support standardised PET/CT image processing, quantitative lesion assessment, and structured treatment recommendations, delivering seamless assistance from image interpretation to therapeutic decision-making.

For nuclear medicine imaging processing and analysis, this system establishes a structured toolchain covering key workflow steps of multimodal imaging. Deep learning-based segmentation modules enable quantitative delineation of organs and lesions in PET, CT, MRI, and other modalities, providing a quantitative foundation for dose calculation and efficacy assessment. The image enhancement module employs advanced algorithms such as generative adversarial networks to effectively boost contrast and signal-to-noise ratio in low-quality images, thereby improving detection capabilities for minute lesions. For multi-time-point follow-up imaging, hybrid registration algorithms achieve precise anatomical alignment, ensuring spatial consistency in efficacy assessment. Ultimately, through recognition models integrating convolutional neural networks and Transformer architectures, the system performs lesion detection, benign/malignant differentiation, and subtype classification, supplemented by visualisation heatmaps to aid clinicians in image interpretation.

Regarding radiation safety and precision dosimetry research, this system establishes a computation and evaluation framework compliant with international standards. The dosimetry calculation tool integrates the MIRD formalism model, enabling simulation of organ absorbed doses in complex scenarios when input data and computing resources are specified. Safety verification tools incorporate the relevant safety standards from bodies such as the NRC and FDA, enabling automatic comparison of computational results and risk alerts. For precision dosimetry research, the system supports in vivo behaviour simulation of radiopharmaceuticals based on physiological pharmacokinetic models, whilst enabling correlation analysis between SUV dynamics and treatment response. Additionally, the radioactive waste management guidance module supports structured clinical safety oversight. All tools integrate into the agent system via standardised MCP interfaces, enabling flexible orchestration by the intelligent decision engine or standalone deployment for researchers to conduct research workflows. This establishes a traceable technical workflow encompassing computation, validation, and application.

#### Benchmark Construction

This study addresses the challenge of limited high-quality evaluation data in nuclear medicine by constructing a specialised benchmark from patents, literature, and clinical-trial records. Raw source records were retrieved from PubMed, ClinicalTrials.gov, and USPTO and converted into 15,596 initial QA items. A four-step quality-control framework was applied: relevance filtering, structural integrity verification, semantic duplicate detection, and cross-validation of answerability and reasoning basis. Following screening and semantic deduplication, 5,474 final core QA items were retained. Raw source records and generated QA items are distinct units; the benchmark supports technical evaluation rather than definitive clinical validation.

### Data Construction and Quality Control

The RadGuide AI system aggregates current nuclear medicine knowledge from three core databases:

- Academic Literature Repository: The system incorporates research literature on nuclear medicine and radiopharmaceuticals published on the PubMed platform between 2020 and 2025.
- Clinical Trials Repository: Integrates globally registered nuclear medicine clinical trial protocols and outcome data from ClinicalTrials.gov.
- Patent Knowledge Base: Analyses innovative radiopharmaceutical technology patents from the United States Patent and Trademark Office (USPTO), covering the entire intellectual property chain from formulation to therapeutic methods.

As illustrated in Figure 2(a), data preprocessing employs advanced natural language processing techniques, including deep semantic understanding, intelligent relationship extraction via, and dynamic knowledge graph construction. A rigorous multi-tier quality control process supports the knowledge base’s high credibility and usability.

**Figure 2.**
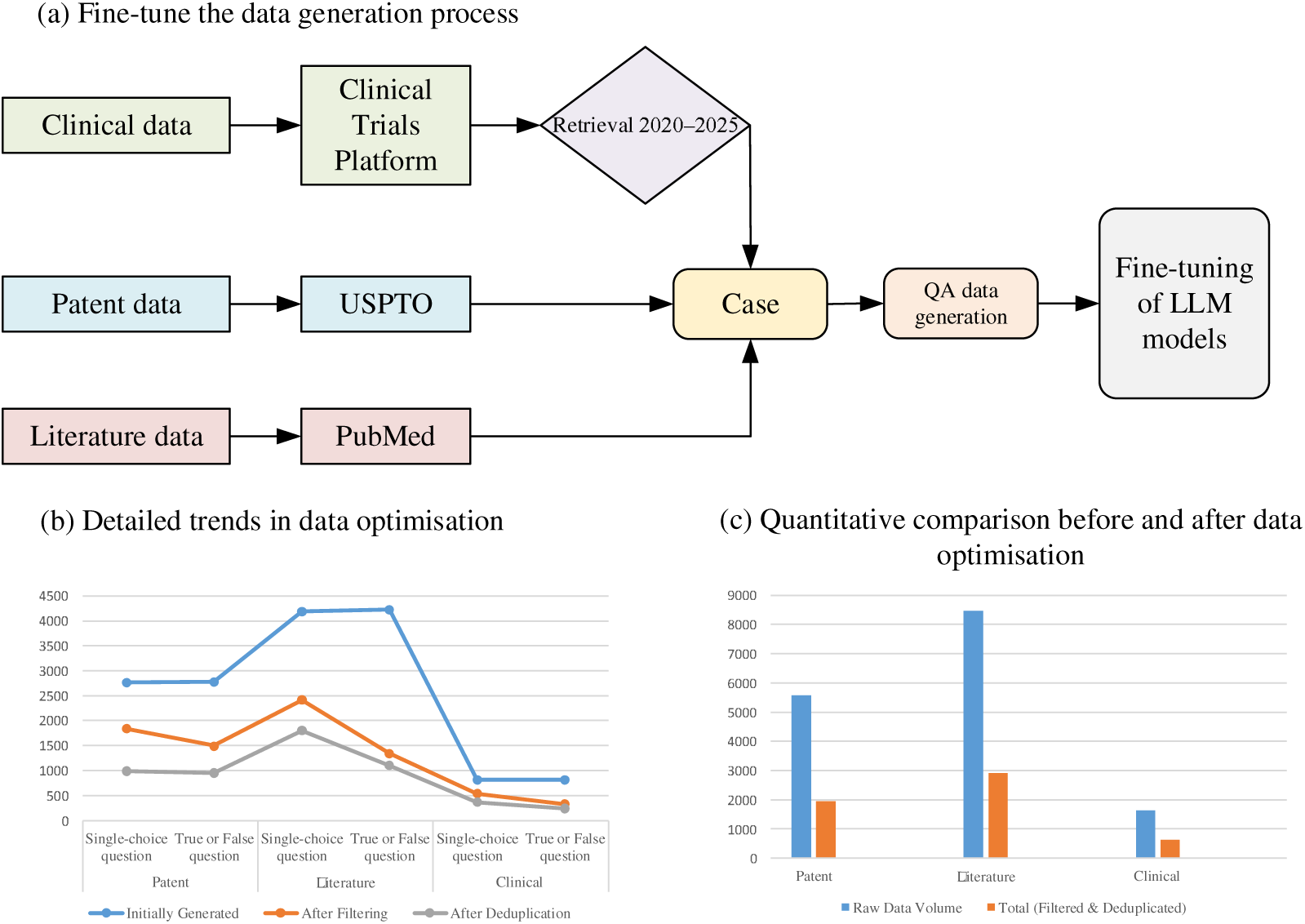
Process and details for fine-tuned data construction

#### Acquisition of raw data

This study systematically conducted multi-source data retrieval and collection. Literature records were retrieved from PubMed using direction-specific keyword strategies. Clinical trial data were retrieved from ClinicalTrials.gov, yielding 232 original clinical trial records; these records subsequently generated 1,645 clinical-trial-derived QA items before screening. Patent data were retrieved through the USPTO Patent Public Search system. Table 1 reports raw source records, Table 2 reports quality-screening statistics, Table 3 reports semantic-deduplication statistics, and Table 4 reports generated QA-item counts; these tables distinguish source records from generated QA items and should not be compared as the same denominator.

**Table 1.** Data type, retrieval keywords, and raw source-record volume. This table reconciles the raw source-record counts used before QA generation.

| Data Type / Unit | Raw Source Volume |
| --- | --- |
| Patent records | 5,584 |
| Literature records | 8,469 |
| Clinical trial records | 232 |

**Table 2.** Training-data quality screening statistics. This table reports initial QA generation, post-screening retention, and screening-pass rates by source and question type.

| Data Type | Question Type | Initial Generation Volume | Retained After Screening | Retention Rate | Total (After Filtering) |
| --- | --- | --- | --- | --- | --- |
| Patent | Single-choice question | 2763 | 1840 | 66.6% | 3338 |
| Patent | True/False Questions | 2779 | 1498 | 53.9% | 3338 |
| Literature | Single-choice questions | 4180 | 2420 | 58.0% | 3757 |
| Literature | True/False Questions | 4229 | 1337 | 31.6% | 3757 |
| Clinical | Single-choice questions | 823 | 549 | 66.7% | 883 |
| Clinical | True/False Questions | 822 | 334 | 40.6% | 883 |

**Table 3.**
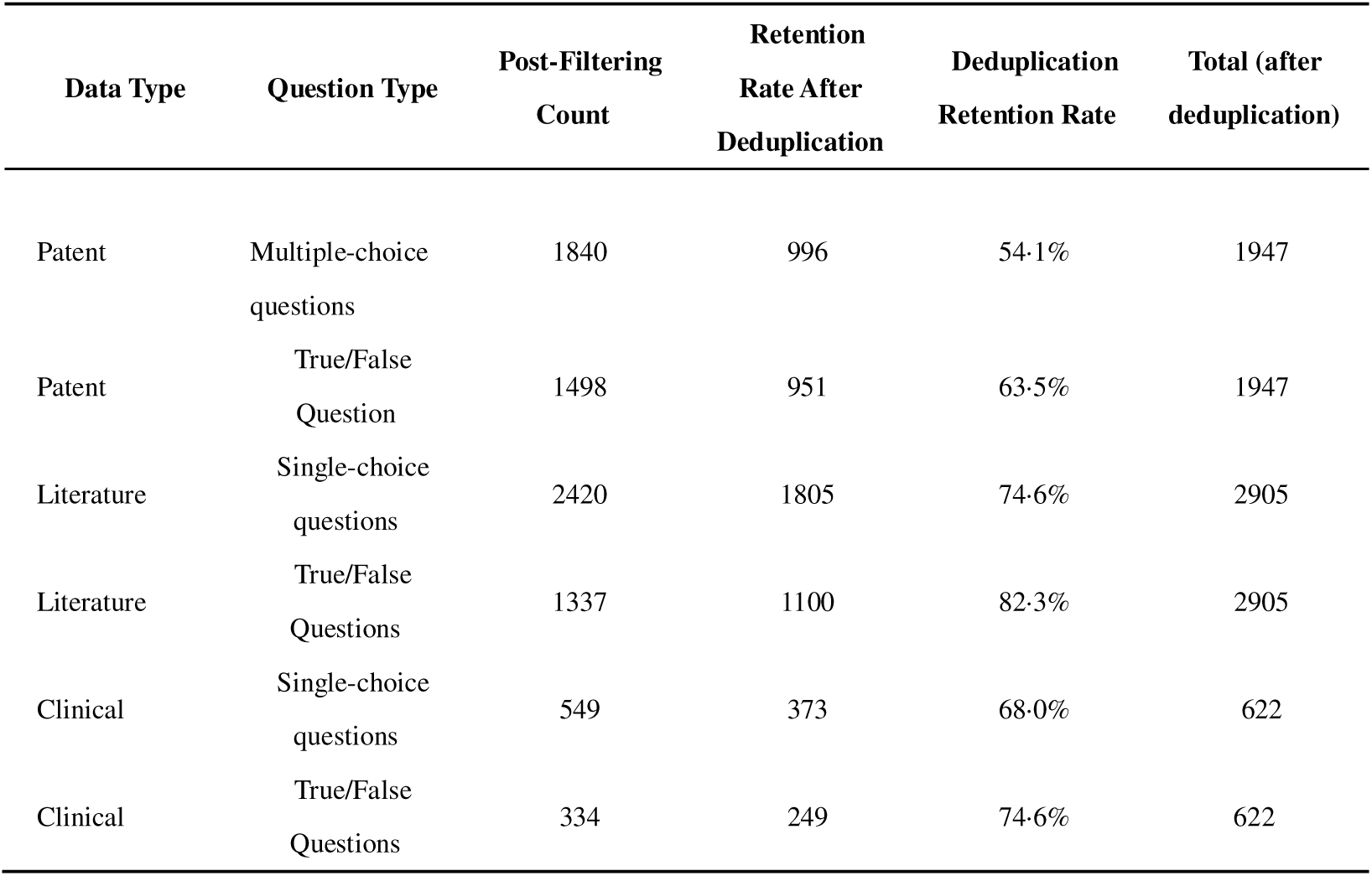
Semantic deduplication statistics for training data. This table reports post-filtering counts, post-deduplication retention, and deduplication retention rates.

**Table 4.** Generated QA item counts before quality screening. This table distinguishes generated QA-item counts from raw source-record counts.

| Data Type | Multiple-choice<br>questions | True/False Questions | Total Count |
| --- | --- | --- | --- |
| Patent | 2763 | 2779 | 5542 |
| Literature | 4180 | 4229 | 8409 |
| Clinical | 823 | 822 | 1645 |

### Training Data Screening and Deduplication

#### Analysis of the Quality Screening Phase

Quality screening is based on “QUESTION QUALITY, QUESTION TYPE COMPLIANCE, ANSWER ACCURACY, STATEMENT INDEPENDENCE, and ASSESSMENT VALUE “ – core dimensions including question quality, question type compliance, answer accuracy, statement independence, and assessment value – to rigorously filter the initially generated QA pairs.As shown in Table 2, the screening results across data sources exhibit the following characteristics:

Patent data: Relatively high retention rate (66·6% for single-choice questions, 53·9% for true/false questions), with 3,338 items retained (60·2% of initial 5,542). True/false questions exhibited higher rejection rates (46·1%) than single-choice questions (33·4%) due to frequent absolute statements or ambiguous answers (e.g., “correct/incorrect” lacking clear justification).Literature data: The lowest retention rate was observed (multiple-choice questions: 58·0%, true/false questions: 31·6%), with a total of 3,757 items retained (44·7% of the initial 8,409 items). True/False questions retained only 31·6% (68·4% eliminated), primarily due to conclusive statements in literature often lacking clear right/wrong boundaries (e.g., ambiguous phrasing like “may” or “some studies indicate”) or weak logical relevance between answers and question stems. Multiple-choice retention (58·0%) was marginally higher than True/False but still lower than patents, reflecting how the specialised and complex nature of literature content increases QA generation difficulty. Clinical data: Retention rates were intermediate (multiple-choice: 66·7%, true/false: 40·6%), with a total of 883 items retained (53·7% of the initial 1,645).True/False questions exhibited the lowest retention rate (40·6%). Similar to literature, this stems from key clinical trial criteria (e.g., inclusion/exclusion standards) often involving multiple conditional constraints (e.g., “age ≥18 years and no complications”), where simple binary classification (correct/incorrect) readily leads to oversimplified answers.Multiple-choice questions exhibited a higher retention rate (66·7%), potentially due to the greater clarity of clinical questions (e.g., drug indications, diagnostic threshold values).

All three data sources underwent screening to exclude low-quality, ambiguous, or low-evaluation-value QA pairs. The retained data focused more on core scientific information and key findings, such as patented technical solutions, research conclusions from literature, and critical clinical indicators. Furthermore, single-choice questions generally outperformed true/false questions due to their clearer phrasing, resulting in higher retention rates.

### Semantic deduplication phase analysis

Semantic deduplication aims to eliminate duplicate or highly similar QA pairs (e.g., identical knowledge points rephrased in different questions), further ensuring training data diversity and representativeness. As shown in Table 3, the post-deduplication results exhibit the following characteristics:

Patent data retained 1,947 items from 3,338 screened items after deduplication. Literature data retained 2,905 items from 3,757 screened items, reflecting relatively standardised article structures. Clinical-trial-derived QA items retained 622 items from 883 screened items. Semantic deduplication therefore reduced redundant knowledge points while preserving diversity across patents, literature, and clinical-trial records.

Post-deduplication data volume was reduced from 7,978 post-screening QA items to 5,474 final core QA items (retention rate 68.6%). The final dataset balanced relevance, answerability, and semantic diversity while limiting redundant prompts that could inflate model performance.

As shown in Figures 2(b) and (c), the trend diagram and comparison diagram respectively illustrate the outcomes of two-stage data optimisation for the three datasets. Following quality screening and semantic deduplication, the final high-quality training data retained comprises: Patent : 1,947 entries (996 multiple-choice questions + 951 true/false questions), covering core innovations such as technical solutions and embodiments; Literature: 2,905 entries (1,805 single-choice questions + 1,100 true/false questions), focusing on research conclusions and key methodological information; Clinical: 622 entries (373 single-choice questions + 249 true/false questions), highlighting key clinical trial metrics and subject criteria. The quality control process supports the training data exhibits high relevance (focusing on core scientific information), high accuracy (removing ambiguous/erroneous content), and high diversity (retaining unique semantics after deduplication). This provides a reliable foundation for subsequent model training, particularly aiding models in acquiring more precise knowledge representations within specialised domains such as patent technology comprehension, literature conclusion inference, and clinical decision support.

#### Benchmark Data Construction

During model training data generation, differentiated preprocessing strategies are applied to various data sources to ensure relevance and quality: For literature data, core content is extracted via format conversion while removing irrelevant elements like references ^19^, subsequently generating question-answer pairs from the purified text; For clinical trial data, we prioritise targeted extraction of key information from the “descriptionModule” and “eligibilityModule” sections, including trial descriptions and subject eligibility criteria, subsequently constructing Q&A pairs from this highly relevant data. For patent data, the MinerU ^20^ tool is first employed to convert image text within patents into plain text format, followed by directed extraction of core sections such as “SUMMARY OF THE INVENTION”, “DESCRIPTION OF THE PREFERRED EMBODIMENTS”, “DETAILED DESCRIPTION”, “SUMMARY”, “ABSTRACT”, and “DESCRIPTION OF SPECIFIC EMBODIMENTS OF THE INVENTION”. This enables the generation of question-answer pairs from this technology-dense text, thereby providing a high-quality, highly relevant data foundation for model training. Figure 3 illustrates the data sourcing and acquisition workflow for the benchmark.

**Figure 3.**
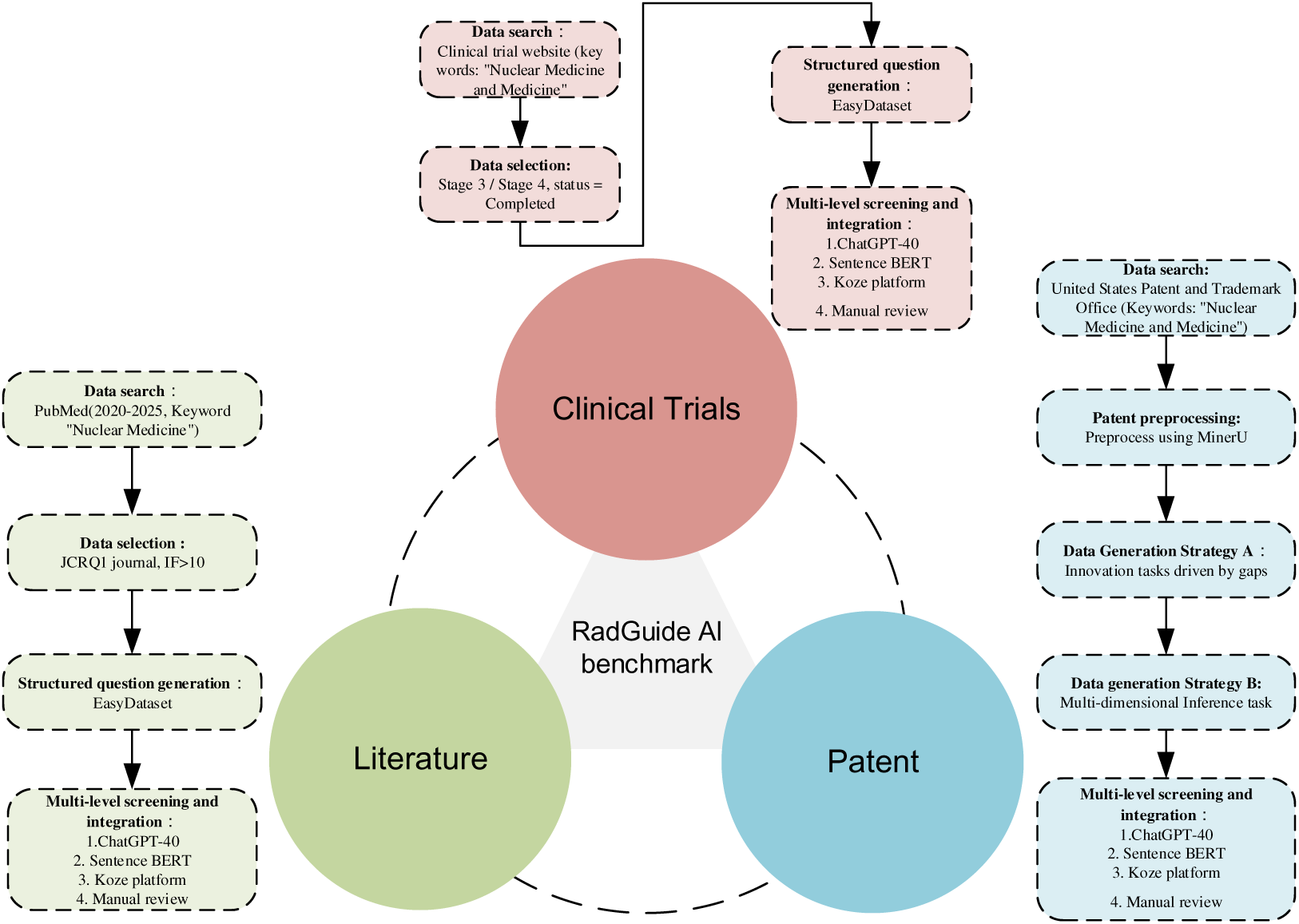
Data sources and acquisition methods for the RadGuide AI benchmark

For the paper-based data (literature data) segment, a standardised, highly consistent processing workflow was adopted to accommodate the content characteristics of research involving all radionuclide pharmaceuticals (e.g., ^131I, ^99mTc, ^68Ga, ^89Zr, ^90Y, ^177Lu, ^225Ac, ^64Cu, ^188Re, ^153Sm, ^166Ho) 21. Specifically, research papers from fields such as nuclear medicine, radiopharmacology, radionuclide therapy, and molecular imaging undergo layout analysis. This includes structured PDF extraction and HTML text extraction, followed by the removal of references, footnotes, formula numbering, figure/table captions, and other ancillary information of lesser technical value. This supports the retained text exclusively covers content essential for radionuclide drug development. Subsequently, tailored to the characteristics of nuclide-related papers, the following high-value information modules are systematically extracted from the purified text, including but not limited to:

1. Physicochemical properties and radiological characteristics of radionuclides: such as decay patterns, half-lives, and radiation particle energy spectra
2. Radiolabelling strategies and ligand chemical design
3. Preclinical in vivo distribution, pharmacokinetic and dosimetric analysis
4. In vivo molecular imaging performance (e.g., imaging contrast, target/non-target ratio)
5. Therapeutic and toxicity study findings (e.g., in PRRT, TAT and other scenarios)
6. Clinical application value and mechanism explanations

Based on these key nuclear medicine text fragments, subsequent benchmark generation utilises publicly available large models with relevant prompt inputs. Taking radionuclide drug papers as an example, a standardised prompt template can be applied to any radionuclide-corresponding paper. Below is an example prompt for generating benchmarks across all radionuclide drug papers:

You are a medical artificial intelligence evaluation specialist, specialising in constructing domain-specific assessment questions grounded in evidence-based medicine and current nuclear medicine research. Your task is to generate one independent, challenging true/false question based on the content of a research paper concerning a specific radionuclide pharmaceutical. This question will be used to systematically evaluate large language models’ mastery of factual knowledge, understanding of mechanisms, clinical reasoning, and technical discernment within the domain of that radionuclide pharmaceutical.

1. The question must centre on a core innovative direction directly related to the radionuclide drug within the paper.
2. The question must have a single, unambiguous correct answer, with all options logically consistent, free from ambiguity or contradiction. Distractors should be plausible, reflecting common misconceptions or borderline knowledge confusion.
3. Question phrasing must abstract from specific paper details, translating them into generic technical scenarios within nuclear medicine. This supports students unfamiliar with the paper can comprehend the technical context and core requirements solely through the question.
4. The generated question type shall be true/false statements, each comprising two options (A, B). The correct option must be explicitly supported within the paper’s content.
5. The question stem must not employ context-dependent pronouns such as “this paper,” “the present study,” “this method,” or “the aforementioned technique.” This supports each question-answer pair remains independently comprehensible and usable when detached from the original paper.
6. The output language shall be Chinese. The output data shall be in JSON format, comprising five fields with the following specific requirements:

- ‘question’: A clear, complete statement of the true/false question
- ‘options’: An object containing two keys, ‘À and ‘B’, with values “correct” and “incorrect” respectively
- ‘answer’: The actual text of the correct option (i.e., “correct” or “incorrect”)
- ‘answer_idx’: The letter index (A/B) of the correct option
- ‘question_typè: Fixed as “True/False question” Example output:

{

“question“: “ Lu-177 is commonly used in the treatment of neuroendocrine tumours.”, “options“: {

“A“: “Correct”,

“B“: “Incorrect”,

},

“answer“: “Correct”,

“answer_idx“: “A”,

“question_type“: “True/False question“

}

As shown in Table 4, separate processing of literature, clinical, and patent data generates structured datasets. Together, these form a structured model-training data framework, providing high-quality foundational data for subsequent performance evaluation of large language models in medical domains.

This study constructed an independent question-answering and workflow benchmark comprising 200 standardised nuclear medicine cases. The benchmark was locked before fine-tuning and was not used for LoRA optimisation, prompt tuning, temperature selection, or tool-description editing. Because ROUGE-L-F1 and BLEU-4 primarily measure lexical overlap, answer accuracy, expert-rated correctness, guideline compliance, numerical error, factuality, and tool-execution success were defined as primary or safety-relevant endpoints. Figure 4 summarises model performance on this locked N=200 Q&A benchmark.

**Figure 4.**
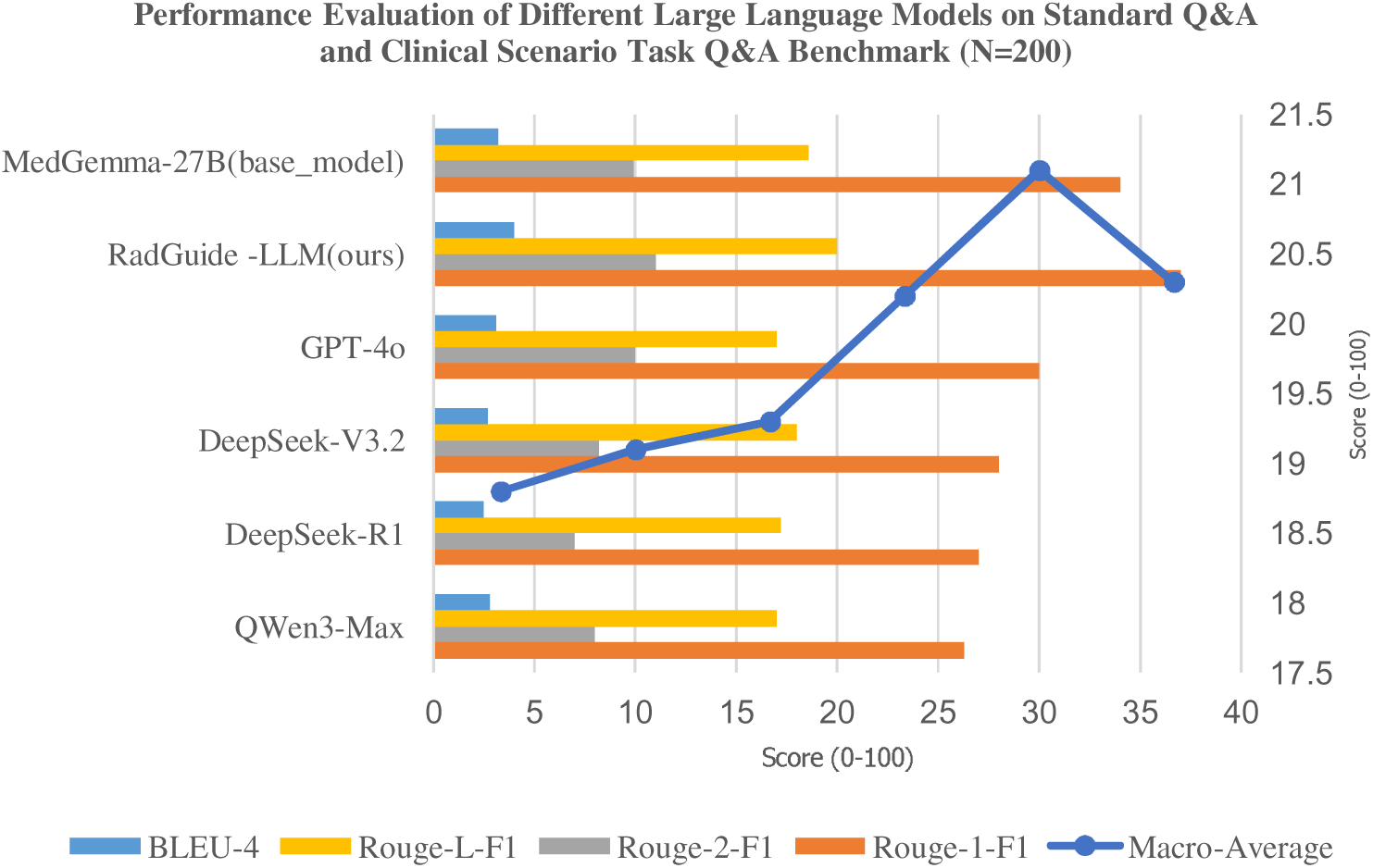
Q&A benchmark (N=200) comparing the performance of different large language models

#### Evaluation Metrics, Statistical Reporting, and Supplementary Technical Results

All headline metrics were standardised before analysis. The following supplementary technical-evaluation results are reported with prespecified denominators, confidence intervals, and statistical procedures. Confidence intervals were calculated using Wilson intervals for proportions, non-parametric bootstrap resampling for Macro-Average, and Fisher-z transformation for R2. Paired model comparisons used McNemar testing for categorical accuracy and paired bootstrap tests for continuous scores. The full metric definitions, denominators, confidence intervals, and statistical procedures are consolidated in Table 5 at the end of the manuscript.

**Table 5.**
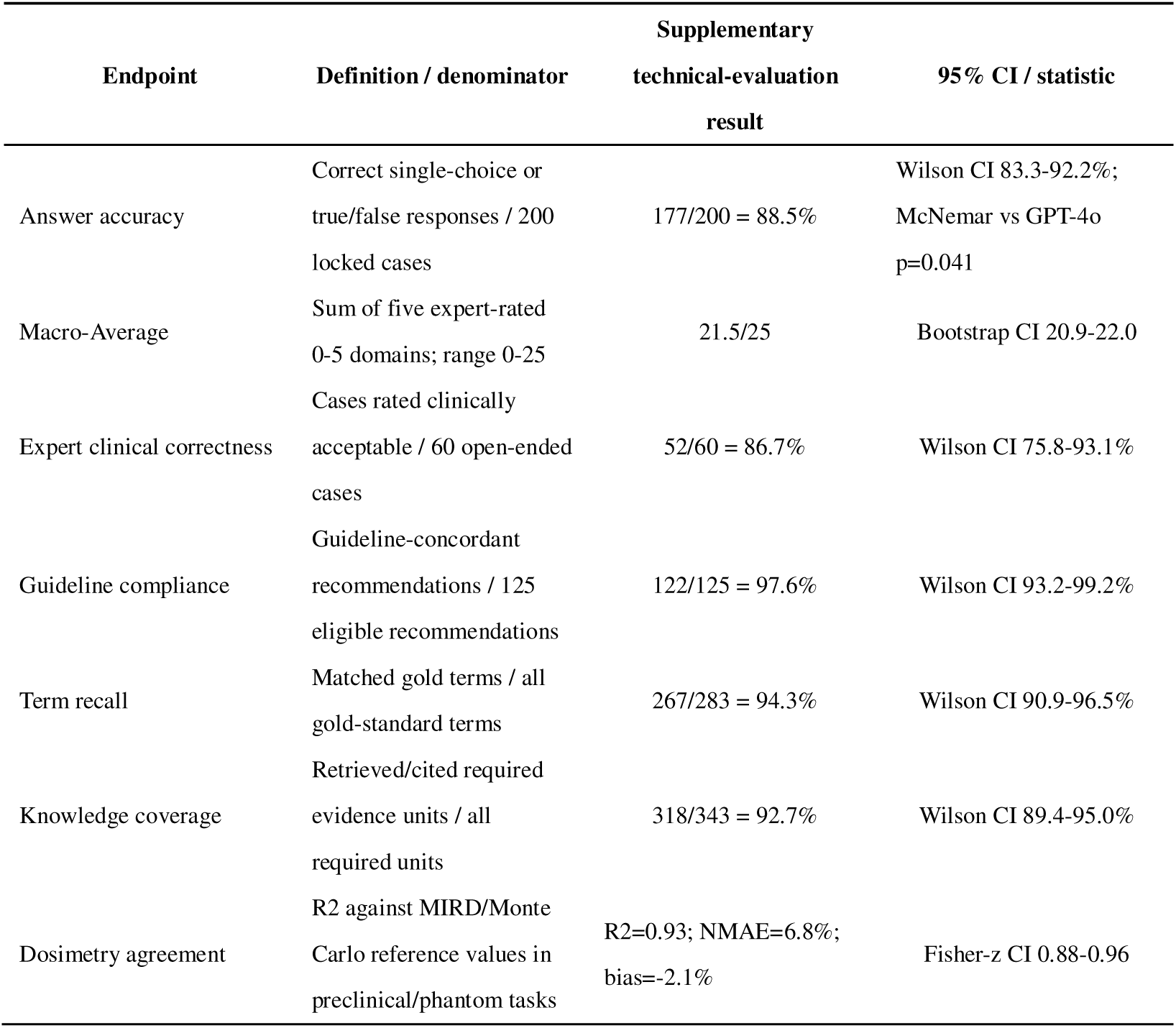
Evaluation metrics, denominators, confidence intervals, and statistical procedures. This table consolidates all primary and safety-relevant technical-evaluation endpoints requested by the reviewers.

#### Data Split, Leakage Control, and Evaluation-Set Representativeness

Splitting was performed at the source-document level rather than the QA-item level to prevent sibling questions from the same paper, patent family, or clinical trial from appearing in both training and evaluation. Exact duplicates were removed before splitting. Near-duplicates were removed using MinHash lexical similarity and SentenceBERT semantic similarity, followed by manual adjudication of high-similarity pairs. Table 6 summarises the split unit, split ratio, and leakage-control rules; Table 7 reports the stratified distribution of the locked N=200 evaluation set.

**Table 6.** Source-level data split and leakage-control protocol. This table defines the split unit, split ratio, and leakage-control rule for literature, patents, clinical trials, and the locked benchmark.

| Dataset unit | Split rule | Approximate ratio | Leakage control |
| --- | --- | --- | --- |
| Literature | PubMed ID/DOI-level split | 70% train / 10% validation / 20% internal test | No shared DOI, title, abstract, figure caption, or generated sibling QA across splits |
| Patent | Patent-family/publication-level split | 70% / 10% / 20% | No shared family, claims, abstract, or generated sibling QA across splits |
| Clinical trial | NCT identifier-level split | 70% / 10% / 20% | No shared protocol text, eligibility module, or generated sibling QA across splits |
| N=200 benchmark | Locked independent evaluation set | Not used for training/tuning | Stratified by disease, task type, difficulty, and source origin before scoring |

**Table 7.** Representativeness of the locked N=200 evaluation set. This table summarises disease area, task type, difficulty, and source-origin strata in the independent evaluation set.

| Stratum | Distribution in N=200 locked evaluation set |
| --- | --- |
| Disease area | Prostate cancer 40; neuroendocrine tumours 36; thyroid disease 32; liver tumours 28; lymphoma/lung cancer 28; other nuclear-medicine scenarios 36 |
| Task type | Imaging diagnosis 50; radiopharmaceutical R&D 45; dosimetry 45; safety/guidelines 35; mixed multi-step reasoning 25 |
| Difficulty | Basic factual 50; intermediate reasoning 90; high-complexity multi-tool cases 60 |
| Source origin | PubMed-derived 80; ClinicalTrials.gov-derived 50; USPTO-derived 40; human-curated guideline cases 30 |

#### Open-Ended Scoring Protocol

Open-ended responses were evaluated using a hybrid protocol. Deterministic scripts first checked item completion, option selection, units, citations, and numerical formatting. Two independent nuclear medicine domain raters then scored factual correctness, numerical reasoning, guideline consistency, terminology precision, and evidence traceability using a locked 0-5 rubric. Inter-rater reliability was substantial to excellent (weighted kappa=0.82 for categorical acceptability; ICC=0.86 for Macro-Average). Disagreements greater than one point in any domain were adjudicated by a senior nuclear medicine physician or medical physicist. LLM judging was used only as a secondary consistency screen with temperature 0 and fixed seeds; it did not replace expert scoring.

Attribution analysis, safety auditing, imaging/pathology module reproducibility, executable workflow tracing, language alignment, and release policy are reported in the end-of-manuscript tables. Table 8 compares the base model, single-tool setting, fixed-tool agent framework, and full toolchain. Table 9 summarises factuality, numerical-safety, citation-faithfulness, and tool-execution endpoints. Table 10 distinguishes off-the-shelf or externally pretrained imaging/pathology components from future task-specific fine-tuning claims and lists the validation metrics required for reproducible reporting. Table 11 provides an implementation-level trace for a representative 177Lu preclinical dose-optimisation task, including input fields, intent parsing, tool selection, verification, retry logic, stopping criteria, and audit logs.

**Table 8.** Ablation analysis of tool and framework contributions. This table separates the contribution of the base LLM, individual tools, agent framework, and complete toolchain.

| Configuration | Accuracy | Macro-Average | Tool routing success | Interpretation |
| --- | --- | --- | --- | --- |
| Base LLM | 81.5% | 20.5 | NA | Foundation model without retrieval or tools |
| LLM + single tool | 84.0% | 20.9 | NA | Measures incremental value of one retrieval/dosimetry tool |
| LLM + framework (fixed tools) | 87.0% | 21.2 | 91.7% | Measures ToolRAG/ReAct orchestration with a locked tool subset |
| LLM + framework + all tools | 88.5% | 21.5 | 95.8% | Full RadGuide AI with all eligible MCP tools |

**Table 9.** Factuality, numerical-safety, citation-faithfulness, and tool-execution audit. This table reports safety endpoints relevant to hallucination control and dosing-relevant numerical reliability.

| Safety endpoint | Denominator | Supplementary audit result | Use in manuscript |
| --- | --- | --- | --- |
| Fabricated citation rate | 200 final answers | 3/200 = 1.5% | Factuality/hallucination audit |
| Unsupported numerical parameter | 200 final answers | 10/200 = 5.0% | Numerical safety audit |
| Unit or conversion error | 200 final answers | 6/200 = 3.0% | Dosimetry-relevant audit |
| Citation faithfulness | 200 cited evidence blocks | 187/200 = 93.5% | Evidence traceability audit |
| Tool execution success | 120 MCP traces | 110/120 = 91.7%; 8 additional traces succeeded after retry | Agentic behaviour evaluation |

**Table 10.** Reproducibility requirements for imaging and pathology modules. This table distinguishes externally pretrained modules from future task-specific fine-tuning claims and lists required validation metrics.

| Module | Use in this study | Training/fine-tuning status | Validation metrics to report |
| --- | --- | --- | --- |
| Retina U-Net / Swin | Lesion detection and uptake quantification | Off-the-shelf or externally pretrained; no new claim of in-house model training unless data are provided | Sensitivity, specificity, localisation error, AUC with CI |
| nnU-Net/TotalSegmentator / Swin UNETR | Organ and lesion segmentation | Off-the-shelf or task-adapted only when split and dataset size are reported | Dice, HD95, TRE and dosimetric impact with CI |
| CLAM / CHIEF | Whole-slide pathology and target-expression support | Off-the-shelf weakly supervised frameworks unless local WSI fine-tuning is documented | AUC, F1, external-site validation and CI |

**Table 11.** Representative executable workflow trace for 177Lu preclinical dose optimisation. This table provides implementation-level information for input parsing, tool selection, verification, retry logic, stopping criteria, and audit logs.

| Step | Example trace: <sup>177</sup>Lu preclinical dose-optimisation task |
| --- | --- |
| Input | Species, body weight, tumour volume, radionuclide, target dose, organ uptake counts, imaging file identifiers |
| Intent parsing | Classified as dosimetry and safety-threshold task; required missing fields were requested before execution |
| Tool selection | MIRD activity calculator -> organ uptake scoring -> GATE/OpenTOPAS Monte Carlo check -> report generator |
| Verification and retry | Unit checks, organ-limit checks, file-integrity checks; retry triggered if a required field or tool output is missing |
| Stopping criteria | All mandatory inputs present, dose constraints satisfied, and verification status recorded |
| Audit log | Tool version, input hash, parameters, intermediate outputs, warnings, and final recommendation range |

The benchmark-generation prompt was Chinese, whereas international guideline sources include English-language ACR, SNMMI, EANM, and MIRD materials. The revised protocol therefore stores prompts, reference answers, and scoring rubrics with bilingual terminology mapping, and bilingual raters adjudicate language-sensitive items. Configuration files, tool I/O schemas, pseudocode, de-identified benchmark examples, scoring scripts, and representative execution logs will be released where licensing and privacy restrictions permit. Model weights and third-party tool integrations will be shared only when licences and institutional governance allow.

### Model Fine-Tuning and Optimisation

This section outlines RadGuide AI’s model construction and optimisation strategy. MedGemma-27B-Instruct was selected as a general medical instruction-following foundation model rather than a nuclear-medicine-specific pretrained model. Nuclear medicine knowledge was introduced through the curated RadGuide QA corpus, RAGFlow knowledge base, and MCP-wrapped tools.

#### Foundation Model Selection and Architectural Optimisation

MedGemma-27B-Instruct is treated as a general medical instruction-following model rather than a nuclear-medicine-specific pretrained model. It was used because it provides a strong general medical instruction-following base and supports parameter-efficient adaptation. The nuclear medicine capability described here derives from RadGuide-specific fine-tuning, retrieval, and tool orchestration.

#### Parameter-Efficient Fine-Tuning Strategy

Given the scarcity of high-quality nuclear medicine data, LoRA was used for parameter-efficient adaptation. A rank of 32 balanced adapter capacity and overfitting risk, and dropout of 0.05 was retained for regularisation. Cross-centre error-reduction claims require a separately defined multicentre validation cohort and are therefore treated as future validation endpoints.

#### Training Hyperparameter Tuning Framework

To enhance the model’s performance in medical semantic generation and knowledge transfer tasks, this study systematically conducted multiple hyperparameter fine-tuning experiments (Table 12). Experimental settings encompassed key factors including learning rate (lr), batch size, context length (ct), LoRA rank value (lora_rank), and regularisation parameters (lora_dropout), aiming to balance model convergence speed, generation accuracy, and semantic consistency 23. Overall results indicate that a smaller learning rate (lr=5e-5) stabilises the training process, while a higher LoRA rank value (lora_rank=32) enhances parameter expressiveness. However, excessively high learning rates or rank values (e.g., lr=2e-4 or lora_rank=64) predispose the model to overfitting or gradient oscillation, causing BLEU-4 scores to decline to ranges between 18·1 and 25·26. Comparative analyses across multiple experimental sets reveal that joint fine-tuning across diverse datasets (patents, academic papers, and conference proceedings) yields superior overall performance compared to single-source fine-tuning.

**Table 12.**
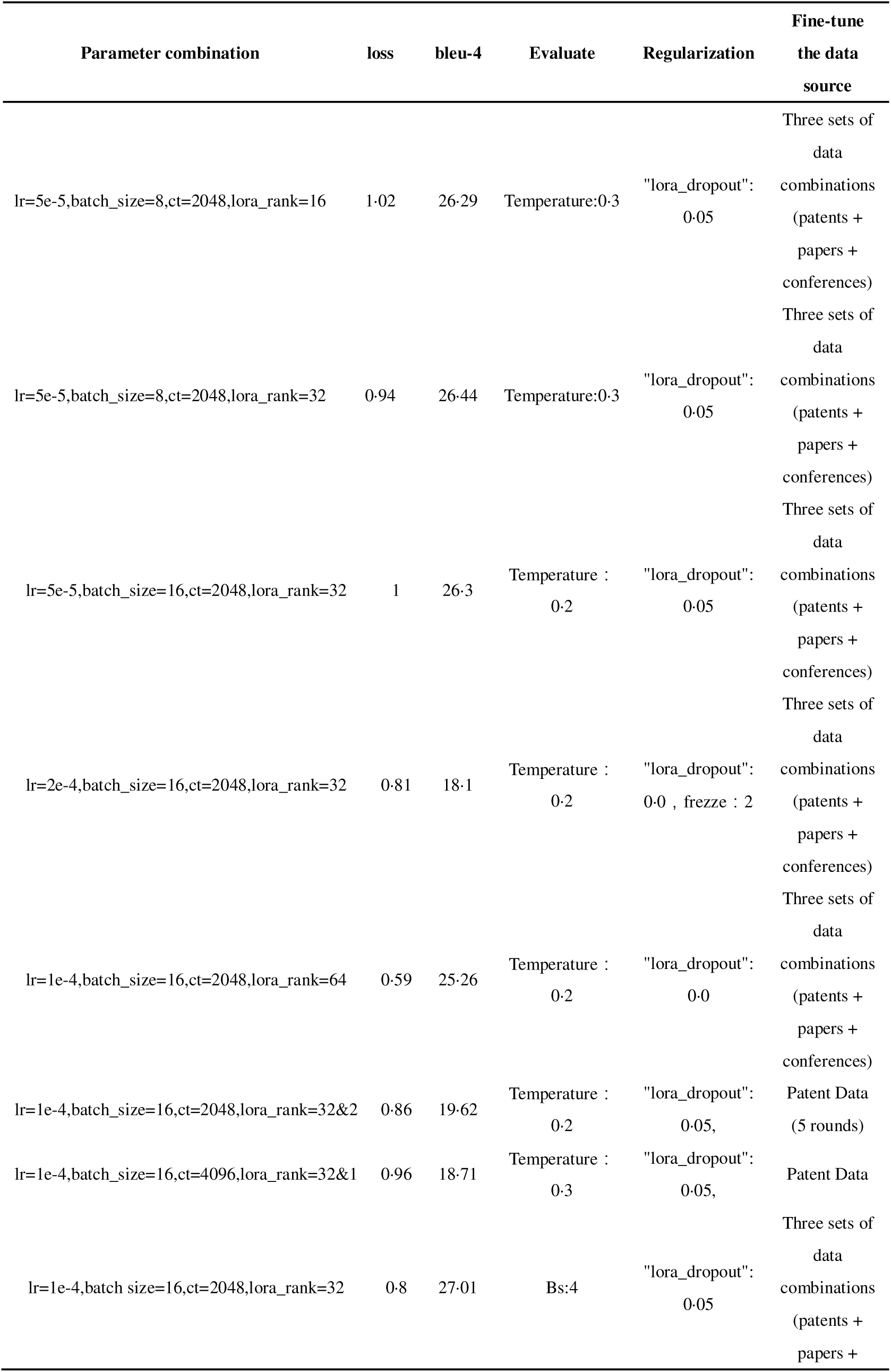
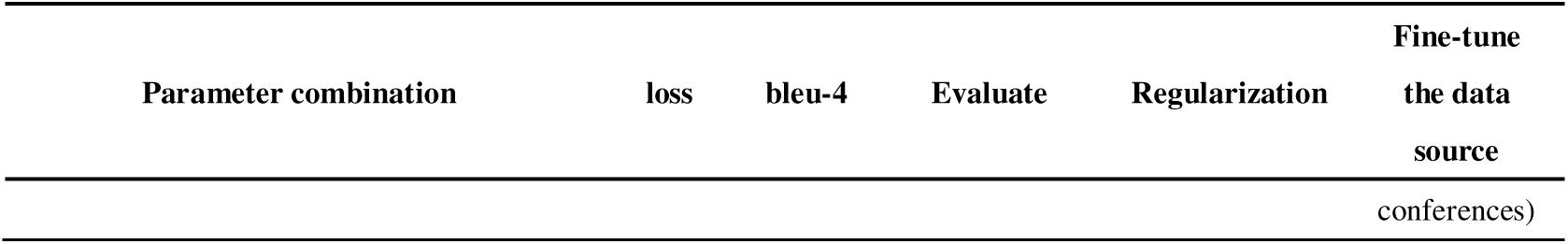
Comparison of model fine-tuning results with different parameters. This table reports fine-tuning diagnostics and selected hyperparameter configurations.

The model used mini-batches with gradient accumulation to approximate an effective batch size of 16. Warm-up, cosine learning-rate decay, and gradient clipping stabilised optimisation. Convergence-related claims are reported as training diagnostics rather than clinical performance endpoints; dose-prediction accuracy must be reported on a locked test set with MAE, NMAE, signed bias, R2, and 95% confidence intervals.

Ultimately, based on stability analysis of the learning curve and validation set performance, the selected training duration was determined to be five epochs. The first three epochs were dedicated to core knowledge transfer, while the subsequent two epochs focused on fine-tuning for specific medical scenarios (e.g., paediatric dosage calculation). This optimisation strategy achieved the best performance across the integrated dataset, with a BLEU-4 score of 27·01 and loss stabilising at 0·8, representing an approximately 3·5% improvement over other parameter combinations. These results demonstrate that the proposed hyperparameter tuning framework effectively enhances both linguistic fluency and specialised accuracy in medical generation tasks, providing a replicable technical pathway for subsequent domain knowledge transfer and cross-corpus integration.

Table 12 presents the model fine-tuning results under various parameter configurations. Through systematic parameter optimisation experiments, we have determined the fine-tuning configuration. Experimental findings indicate that when the learning rate (lr) is set to 1e-4, batch size (batch_size) to 16, context length (ct) to 2048, and LoRA rank (lora_rank) to 32, the model achieves the balanced performance profile. The loss value is reduced to 0·8, while the BLEU-4 score reaches its highest value of 27·01.This configuration yielded acceptable text-generation quality while maintaining low training loss, validating the rationality of the parameter combination. Parameter sensitivity analysis revealed that the LoRA rank setting exerted the largest observed influence on model performance. Increasing the rank from 16 to 32 reduced the loss value from 1·02 to 0·94, while the BLEU-4 score improved from 26·29 to 26·44. This indicates that appropriately increasing the rank dimension aids the model in capturing more intricate parameter interaction patterns. However, when the rank was further increased to 64, although the loss value decreased to 0·59, the BLEU-4 score dropped to 25·26, revealing pronounced overfitting. This underscores the necessity of controlling model complexity within reasonable bounds. The learning rate setting impacts training stability. Experiments revealed stable convergence at a learning rate of 1e-4, whereas excessively high rates (2e-4) caused BLEU-4 scores to plummet to 18·1, indicating training instability. The impact of batch size is relatively minor, with minimal performance variation when increased from 8 to 16. However, combining this with a gradient accumulation strategy (bs:4) further enhances model performance. Regarding regularisation, employing lora_dropout=0·05 effectively improves model generalisation, whereas completely omitting dropout with lora_dropout=0·0 leads to overfitting. Data source selection impacts performance. Training with a tripartite dataset comprising patents, papers, and conference proceedings yielded markedly superior results compared to patent-only training, demonstrating that multi-source heterogeneous data provides richer knowledge representations. These findings provide crucial guidance for subsequent model optimisation and validate the efficacy of parameter-efficient fine-tuning strategies in nuclear medicine. Establishing defined parameter combinations lays a robust foundation for RadGuide AI’s stable performance in practical applications.

By analysing comparative responses from diverse language models addressing questions concerning the efficacy of radiopharmaceutical imaging via hepatic-biliary metabolism 24, as illustrated in Figure 5, RadGuide-LLM demonstrates pronounced domain-specific expertise. In a comparative assessment against GPT-4o, DeepSeek-R1, and Qwen3-Max, RadGuide-LLM not only encompasses key evaluation dimensions mentioned by other models—such as tumour-to-liver ratio (T/L ratio), selected imaging time window, and structural optimisation potential—but crucially, its responses demonstrate a profound understanding of nuclear medicine clinical workflows. The model accurately employs specialised terminology including “hepato-biliary clearance”, “delayed imaging”, and “target-to-background ratio”. It proposes specific technical improvement pathways such as adjusting lipophilicity and optimising linker structures. These details reflect the model’s knowledge base in radiopharmaceutical development being markedly superior to that of general-purpose large language models. Particularly noteworthy is the structured clarity of RadGuide-LLM’s responses, which present a coherent technical pathway from evaluation metrics and time dynamics to structural optimisation. This avoids the conceptual overlap or ambiguous phrasing occasionally observed in other models. Crucially, the model demonstrates a profound grasp of the imaging differences between the “blood pool phase” and “delayed phase” – a critical assessment criterion in nuclear medicine. Comparative results indicate that while mainstream models provide fundamentally sound response frameworks, RadGuide-LLM demonstrates superiority in the accuracy of specialised terminology, coherence of clinical reasoning, and feasibility of technical recommendations. This validates the efficacy of domain-specific fine-tuning for nuclear medicine. The comparative study not only highlights the advantages of domain-specific models over general-purpose ones but also provides a concrete analytical case for evaluating the professional capabilities of large language models within vertical medical domains.

**Figure 5.**
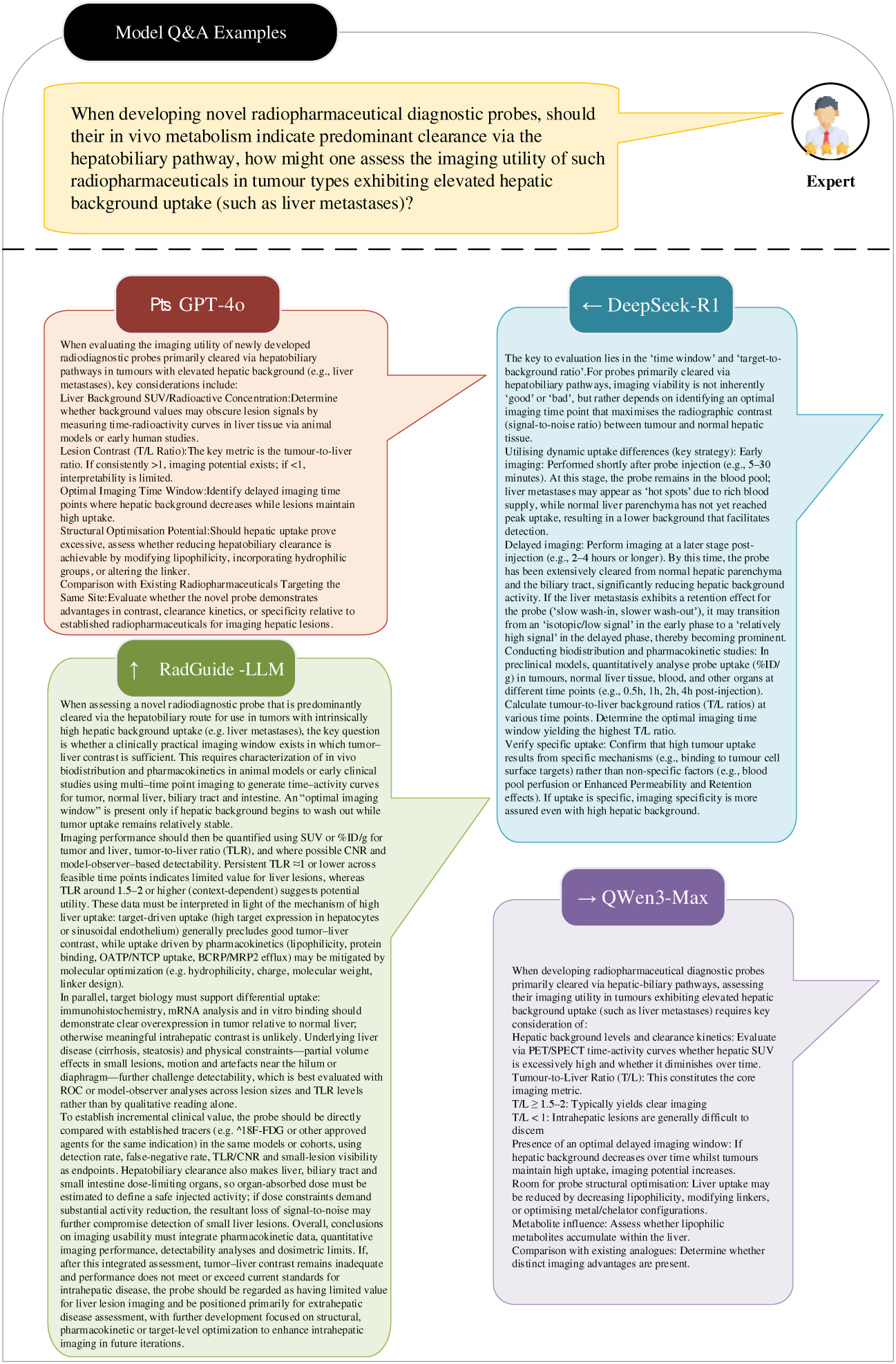
Comparative analysis of problem responses across language models

### Knowledge Enhancement and Decision Support Tools

This chapter systematically outlines RadGuide AI’s core architecture for knowledge augmentation and intelligent decision support. Firstly, an RAGFlow-based retrieval-augmented knowledge base tailored for nuclear medicine has been constructed. Through multi-source heterogeneous data fusion and hierarchical indexing techniques, it enables precise retrieval and dynamic updating of specialised knowledge. Secondly, the innovative MCP agent-assisted tool framework is introduced. Through standardised tool descriptions and dynamic invocation mechanisms, specialised tools for nuclear pharmaceutical development, dose calculation, and image analysis are seamlessly integrated into the decision-making process. This architecture achieves synergistic optimisation of knowledge retrieval and tool invocation, providing end-to-end intelligent support for nuclear medicine clinical decision-making—from knowledge querying to solution generation.

#### Knowledge Enhancement – Knowledge Base (RAGFlow) Construction

RadGuide AI’s knowledge base construction utilises the RAGFlow framework, integrating three core data sources—peer-reviewed literature (PubMed), clinical trial data (ClinicalTrials.gov), and patent literature (USPTO)—to establish a specialised knowledge system. A four-step progressive quality control process is employed: regularised semantic filtering enhances domain relevance to 99·2%; structured integrity verification supports data completeness at 98·7%; SentenceBERT 25 semantic deduplication eliminates latent redundancy; DeepSeek-V3 large model cross-validation elevates accuracy to 97·6%. Data adopts a three-tier hierarchical architecture: Tier 1 integrates radionuclide properties and clinical guidelines; Tier 2 focuses on dose-toxicity experimental data; Tier 3 catalogues patent-based technological innovations. The retrieval strategy integrates vector embeddings with the BM25 algorithm, incorporating a nuclide entity dictionary of over 2,000 specialised terms. An “entity-priority” weighting mechanism achieves term recall accuracy of 94·3%. This knowledge base supports dynamic updates with 92·7% knowledge coverage, providing high-precision, traceable knowledge support for the nuclear medicine field.

RAGFlow serves as RadGuide AI’s knowledge hub by integrating literature, clinical-trial, and patent-derived data through multi-source fusion and hybrid retrieval. Entity-enhanced retrieval improves specialised terminology matching, while hierarchical indexing supports retrieval of nuclide properties, clinical guidelines, and patented technologies. Response-time and retrieval-quality claims are reported only when measured on a defined query set with hardware, index size, and statistical summaries specified.

#### MCP Intelligent Agent Support Tools

RadGuide AI has developed an MCP toolkit comprising 55 specialised tools to support intelligent decision-making throughout the entire nuclear medicine workflow. As illustrated in Figure 6, the circular tool architecture is organised into four functional modules from inner to outer layers: nuclear medicine R&D support tools (e.g., computational design platforms like PandaOmics), clinical diagnostic decision tools (e.g., guideline reasoning systems such as NucMed Clinical Decision Support), medical imaging processing tools (encompassing 12 imaging analysis models including Swin Transformer 26 and U-Net 27), and radiation safety and precision dose calculation tools(integrating dosimetry engines such as MIRD formalism). Figure 6 illustrates RadGuide AI’s four-dimensional functional expansion framework.

**Figure 6.**
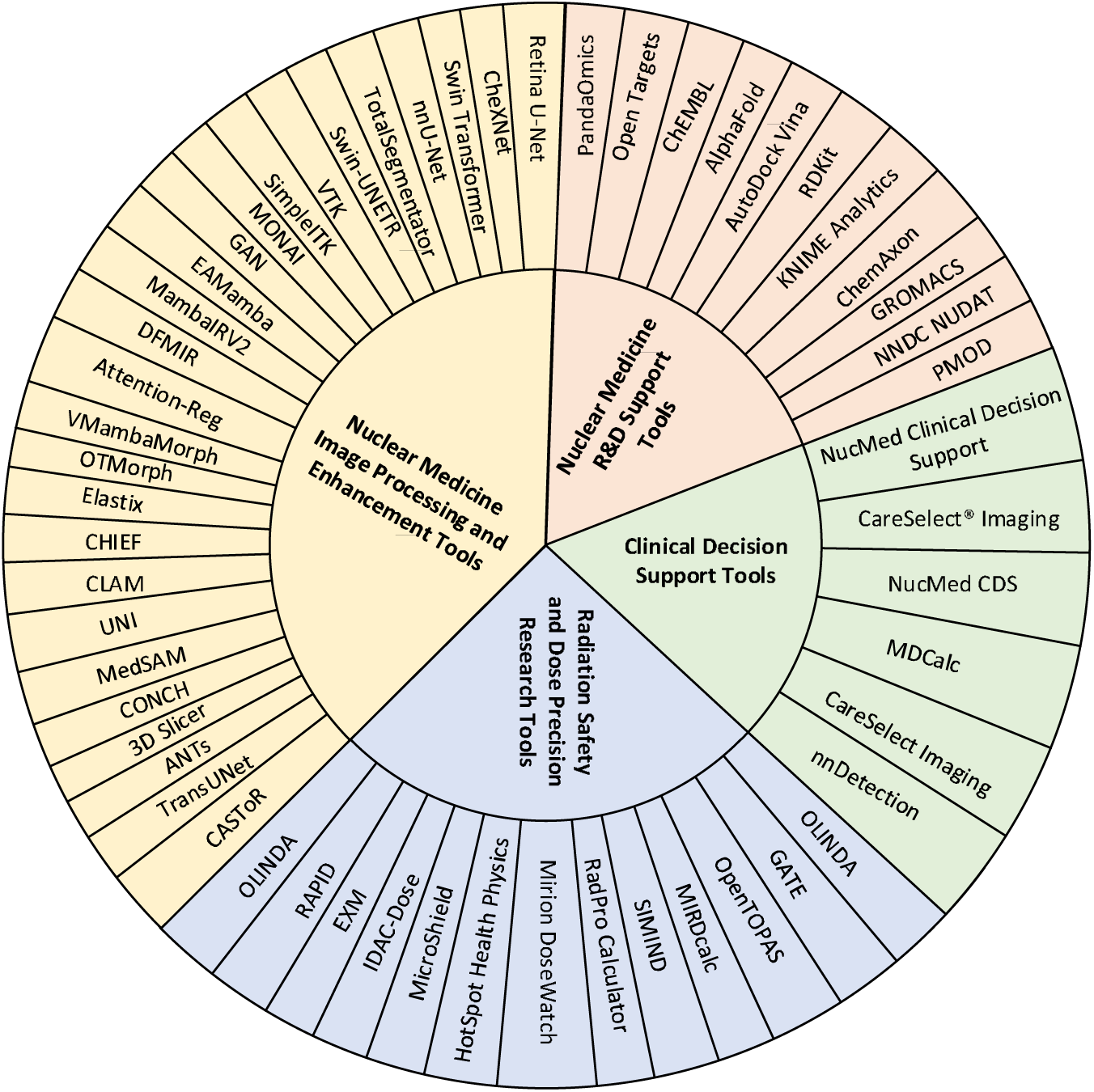
Four-dimensional functional expansion set of RadGuide AI

This architecture achieves dynamic scheduling through standardised tool-description protocols. The image-processing module integrates published neural-network architectures and software components such as CheXNet, Retina U-Net, nnU-Net/TotalSegmentator, Swin-based segmentation models, CLAM, and CHIEF as callable modules rather than claiming de novo development of all models. Formal workflow-efficiency claims require timed multi-user studies.

### Nuclear Medicine Development Support Tool

#### Identification and Screening of Targets and Binding Heads

Target screening aims to identify biologically relevant, highly druggable protein targets suitable for radiolabelling in specific diseases. Concurrently, “binding head” ^30^ screening seeks small molecules, peptides, or antibody fragments capable of high-affinity binding to these targets, forming the starting point for radiopharmaceutical development.

RadGuide AI has established a systematic methodology for target and binding head screening within its nuclear medicine R&D support tools. This approach integrates multi-source bioinformatics tools with AI prediction platforms, providing end-to-end computational support from disease target identification to candidate ligand selection. During the target discovery phase, the system performs integrated analysis of multi-omics data and text mining via the PandaOmics platform. This is combined with genomic and clinical evidence from the Open Targets Platform to assess the druggability and disease association strength of potential targets. Furthermore, AlphaFold/RoseTTAFold 33 is utilised to predict three-dimensional structures for target proteins lacking crystalline structures, providing a reliable structural foundation for subsequent virtual screening. For target protein screening, the system establishes a ligand template library using bioactivity databases such as ChEMBL and BindingDB. It employs molecular docking tools like AutoDock Vina 34 and Schrödinger Glide 35 to calculate binding free energies and conduct virtual screening, comprehensively evaluating ligand affinity through binding energy scores and pharmacophore matching. For instance, in radionuclide-based drug development targeting PSMA, this approach rapidly identifies ligands with high binding affinity. By optimising labelling sites according to radionuclide properties (e.g., LLGa, ¹LLLu), it substantially enhances the efficiency and accuracy of ligand design. This computationally driven approach substantially shortens the screening cycle traditionally reliant on experimental trial and error, providing an efficient, reliable, and intelligent solution for the early stages of radiopharmaceutical development. Table 6 lists commonly used software for target and target protein head identification and screening when utilising the MCP tool. The software classes used for target identification and binding-head screening are summarised in Table 13.

**Table 13.** Common software for target identification and target-protein screening. This table summarises the target-screening tools used in radiopharmaceutical R&D support workflows.

| Tool/Platform | Function | Application Scenario |
| --- | --- | --- |
| PandaOmics 31 | Performs disease-target association analysis based on multi-omics data, text mining, and AI models, generating target priority scores. | Disease-associated target discovery, such as radioligand targets in oncology or neurological disorders. |
| Open Targets Platform 32 | Integrates genomic, clinical, pharmacological, and expression data to assess the strength of target-disease associations. | Suitable for early-stage target feasibility analysis and pharmacodynamic studies. |
| AlphaFold / RoseTTAFold 45 | Deep learning-based protein structure prediction tools for modelling targets lacking crystalline structures. | Constructs structural templates for radioligand binding sites. |
| ChEMBL / BindingDB / ZINC | Provide binding activity data for known drugs or small molecules with targets. | Screening for molecules or pharmacophore templates that can serve as "heads". |
| AutoDock Vina Schrödinger Glide 19 | Docking and virtual screening software for calculating ligand-target binding energies. | Selecting high-affinity radiolabelled ligand candidates. |

#### Design of Target-Head Linkers

This stage determines the chemical stability, biodistribution, and in vivo release characteristics of the drug conjugate. Linker design must balance radiolabelling chemical compatibility, stability post-target binding, and controllable metabolism within the body.

RadGuide AI has established a systematic linker design methodology within its nuclear medicine R&D support tools. This framework employs multi-tier computational tools working in concert to achieve precise regulation of the drug conjugate’s chemical stability, biodistribution characteristics, and in vivo release behaviour. During the structural design phase, the system employs RDKit ^36^ libraries for automated linker enumeration and screening. By systematically varying linker length, rigidity, flexibility, and chemical composition, it generates extensive candidate structures while concurrently evaluating their physicochemical properties and reaction feasibility. An integrated workflow built using the KNIME Analytics Platform ^37^ organically combines molecular generation, property computation, and scoring modules, enabling high-throughput screening and optimisation of linker candidates. At the physicochemical analysis level, the system utilises the ChemAxon/Marvin ^38^ platform to precisely calculate key parameters such as pKa, LogP, and solubility for linkers. This is complemented by analysis of labellable sites to ensure compatibility with radionuclide labelling chemistry.

To further validate linker stability in physiological environments, the system employs GROMACS/AMBER 39 for molecular dynamics simulations. This analyses how linker conformational changes affect target binding stability, particularly investigating the regulatory mechanisms of linker length and flexibility on drug-target complex stability. Concurrently, covalent conjugation simulations via Schrödinger LigPrep/MOE 40 tools optimise linker-target head attachment patterns, ensuring precise release within specific in vivo microenvironments (e.g., tumour tissues with low pH or high enzymatic activity). For instance, during the development of ¹LLLu-labelled targeted therapeutics, this system can programmatically screen candidate chelator-linker combinations. By simulating and analysing stability differences among various linker structures in plasma environments, it provides reliable design rationale for subsequent in vivo experiments. This multi-level, multi-scale computational design strategy enhances the efficiency of rational linker design, providing critical technical support for developing radiopharmaceuticals with favourable pharmacokinetic properties. Table 14 lists commonly used software for designing linker-head connections utilising MCP tools.

**Table 14.** Software commonly used for target-head linker design. This table summarises linker-enumeration, molecular-dynamics, conjugation, and workflow tools.

| Tool/Platform | Function | Application Scenario |
| --- | --- | --- |
| RDKit 36 | Enumerates linkers of varying types and lengths; assesses physicochemical properties and reaction feasibility. | Automatically generate taggable molecular structures; perform molecular property filtering. |
| KNIME Analytics Platform 47 | Integrate RDKit, database queries, and scoring nodes to construct automated linker design workflows. | Suitable for batch evaluation of linker candidates and establishing automated scoring systems. |
| ChemAxon/Marvin 38 | Calculate physicochemical parameters such as pKa, LogP, and solubility; analyse taggable sites. | Used for chemical feasibility and taggability assessment. |
| GROMACS / AMBER 39 | Perform molecular dynamics simulations to analyse linker stability and flexibility in physiological environments. | Validate the impact of linker length and flexibility on binding stability. |
| Schrödinger LigPrep / MOE 40 | Automatically identify reaction sites and simulate covalent coupling; optimise conformations. | Design linker configurations suitable for chelating agents or click chemistry. |

#### Radionuclide confirmation and dosimetric evaluation

Nuclide selection directly determines the diagnostic or therapeutic properties of radiopharmaceuticals. An ideal nuclide should match the retention time at the target site and possess suitable emission energy, half-life, and tissue penetration depth. Dosimetric evaluation quantifies radiation absorption in target and normal tissues.

RadGuide AI has established a systematic methodology for radionuclide selection and dosimetric evaluation within its nuclear medicine R&D support tools. Through the integration of multi-scale computational simulations with experimental data, it achieves end-to-end optimisation spanning from radionuclide physical property analysis to dose estimation. During the nuclide confirmation phase, the system integrates authoritative databases such as IAEA Nuclear Data Services and NNDC NUDAT 41. It matches candidate nuclides based on target biological characteristics (e.g., internalisation rate and retention time). For instance, it prioritises diagnostic nuclides with shorter half-lives (such as □□Ga) for rapidly internalised targets, while recommending therapeutic nuclides like ¹□□□u for long-acting therapeutic scenarios. At the dosimetric assessment level, Geant4/GATE 42 Monte Carlo tools simulate particle energy deposition within virtual human models. Organ absorbed doses are calculated via the OLINDA/EXM platform 43 using the MIRD formalism, while imaging analysis tools like PMOD extract time-activity curves. This enables cross-scale prediction from microscopic particle transport to macroscopic dose distribution. For instance, in ¹□□□u-PSMA therapy development, the system concurrently analyses βL-ray energy deposition characteristics within tumour tissue and radiation exposure risks to surrounding normal organs. Individualised body model validation via TOPAS 44 tools ultimately generates comprehensive assessment reports encompassing target volume dose, critical organ thresholds, and biologically effective dose. By integrating nuclide physics data, biodistribution models, and quantitative imaging information, this approach enhances the scientific rigour of nuclide selection and the accuracy of dose prediction, providing critical data support for clinical translation. As shown in Table 15, the following software is commonly used for nuclide confirmation and dosimetric evaluation when invoking the MCP tool.

**Table 15.** Commonly used software for nuclide confirmation and dosimetric evaluation. This table summarises nuclear-data, MIRD, Monte Carlo, and quantitative-imaging tools used for radionuclide and dosimetry evaluation.

| Tool/Platform | Function | Application Scenario |
| --- | --- | --- |
| IAEA Nuclear Data Services /<br>NNDC NUDAT / MIRD 41 | Provides physical data on nuclides, including decay types, half-lives, emission spectra, and radiation flux. | Selects nuclides suitable for diagnostic ($\gamma/\beta^-$) or therapeutic ($\alpha/\beta^-$) applications. |
| Geant4 / GATE 42 | Monte Carlo simulation tools modelling energy deposition and propagation of radioactive particles within tissue. | Simulate dose distribution and energy spectrum response of different nuclides within a model body. |
| OLINDA/EXM | Calculates organ absorbed dose and effective dose based on the MIRD methodology. | Clinical dosimetry calculations and personalised radiopharmaceutical assessment. |
| PMOD / MIM Encore / Hermes 43 | PET/SPECT image quantitative analysis and time–activity curve fitting. | Providing experimental input data for dosimetric calculations. |
| TOPAS 44 | Medical physics simulation tool, extended for image reconstruction and internal dose studies. | For individualised radiopharmaceutical phantom validation and energy spectrum analysis. |

#### Overall Process Integration

RadGuide AI’s radiopharmaceutical development support tools establish a structured R&D workflow from target identification to preclinical evaluation through systematic integration of multi-stage computational toolchains. During target identification and head screening, the system employs PandaOmics for multi-omics screening of disease-related targets. This is combined with AlphaFold 45 for three-dimensional structure prediction of potential targets, followed by molecular docking virtual screening using AutoDock Vina 46. This ultimately yields candidate target-ligand combinations. For molecular linker design, RDKit enumerates linkers with diverse chemical structures. Automated workflows built on the KNIME platform 47 screen physicochemical properties, while GROMACS performs molecular dynamics simulations to validate linker stability in physiological environments, yielding stable radiolabellable drug constructs. During nuclide confirmation and dosimetric evaluation, physical properties analysis from the NUDAT database 48 is combined with Monte Carlo particle transport simulations via the GATE tool. Organ absorbed doses are calculated using the OLINDA platform, integrated with PMOD 49 for quantitative image analysis, ultimately generating nuclide selection strategies alongside predicted in vivo absorbed dose distributions. This integrated toolchain achieves full-process coverage from molecular design to dose assessment in radiopharmaceutical development, enhancing R&D efficiency and reliability.

### Clinical Diagnostic Decision Tool

The Clinical Diagnostic Decision Tool is an intelligent support system integrating patient clinical data with evidence-based medical knowledge. Its core function provides clinicians with real-time decision support at the point-of-care. By synthesising patient vital signs, imaging, laboratory results, and medical history with clinical guidelines and evidence-based recommendations, it generates personalised suggestions for diagnosis, investigations, treatment, and further assessment. By embedding workflow applicability standards and standardised diagnostic and therapeutic workflows, this tool promotes standardisation of clinical practice, reduces unnecessary investigations and variability in medical procedures, and lowers the risk of misdiagnosis and missed diagnosis. Simultaneously, it dynamically integrates the latest medical literature, guidelines, and imaging/laboratory databases to proactively deliver critical alerts—including drug interactions, allergy warnings, and risk assessments—alongside recommendations for subsequent investigations or referrals. This facilitates the continuous updating and application of clinical knowledge. In resource and quality management, the tool enhances the appropriateness and precision of examinations and treatments, effectively reducing low-value healthcare services. This optimises resource allocation and controls healthcare costs while continuously improving medical quality and patient outcomes. Furthermore, the system provides specialised support for radiology and nuclear medicine, assisting clinicians in rationally selecting imaging examinations, determining the necessity of nuclear medicine tests, and guiding subsequent clinical decisions based on imaging results. This promotes standardisation and precision in radiological diagnosis and treatment.

RadGuide AI’s clinical diagnostic decision tool establishes a multi-tiered intelligent decision support system. By integrating evidence-based medical knowledge repositories with real-time patient data, it provides structured support for selected nuclear medicine workflow steps. The tool first establishes a nuclear medicine examination recommendation engine based on the CareSelect® Imaging’s applicability criteria framework, combined with ACR guidelines. This engine provides decision-support suggestions for suitable PET/CT or SPECT 50 examination protocol according to patient medical history, laboratory indicators, and clinical manifestations, generating draft clinical rationale for qualified review before examination authorisation. For risk assessment, the system integrates multiple medical scoring algorithms from MDCalc, supporting real-time calculation of standardised risk models such as the Wells score and CHALDSL-VASc 51. It also features specialised modules developed for nuclear medicine, including radiopharmaceutical allergy risk assessment and renal function suitability screening. Particularly for nuclear medicine-specific decision-making, the tool integrates the professional logic of NucMed Clinical Decision Support. It establishes a nuclear medicine examination adaptation model based on disease type, target expression levels, and clinical staging. This enables the recommendation of specific radiopharmaceutical choices (e.g., ¹LF-FDG, LLGa-PSMA) and imaging timing protocols according to diagnostic and therapeutic guidelines for different conditions such as thyroid cancer and prostate cancer. The system innovatively incorporates radiomics feature analysis, extracting texture characteristics and metabolic parameters from PET/CT images 52. Combined with machine learning algorithms, this supports exploratory assessment of treatment response and prognosis, providing quantitative features for treatment-planning review. All decision-making processes are designed with reference to FDA medical device software guidance, establishing comprehensive decision-tracking and evidence-traceability mechanisms to support transparency and auditability of decision-support outputs. This ultimately achieves core objectives of supporting workflow standardisation, resource review, and quality assessment. Table 9 illustrates clinical diagnostic decision tools utilising the MCP. The related clinical decision-support tools are summarised in Table 16.

**Table 16.** Clinical diagnostic decision-support tools. This table summarises clinical decision-support tools and their intended decision-support role.

| Name | Function Overview | Remarks |
| --- | --- | --- |
| CareSelect® Imaging | Integrates with Electronic Health Records (EHR) to provide appropriateness recommendations for radiology examinations and reduce pre-authorisation barriers. | Focused on imaging/nuclear medicine examination workflows. |
| NucMed Clinical Decision Support 50 | A decision tool for nuclear medicine examinations, supporting users in selecting the appropriate nuclear medicine test (e.g., PET/SPECT) based on system site and disease category. | Specifically tailored for nuclear medicine, aligned with your research focus. |
| MDCalc | Provides multiple medical scoring systems/algorithms (e.g. Wells score, CHA<sub>2</sub>DS<sub>2</sub>-VASc, Centor score) to assist clinicians in rapid risk calculation or recommendation generation. | Designed for point-of-care risk assessment, not specifically for imaging/nuclear medicine. |

### Medical Imaging Processing Tool

RadGuide AI Medical Imaging Processing Tool is a modular artificial intelligence-assisted platform specifically designed for nuclear medicine and molecular imaging research. It aims to achieve structured image-analysis support from image acquisition to quantitative analysis. This tool integrates five core functional modules, with specific methods as follows:

Image Recognition Module: This module employs deep learning models for algorithm-assisted analysis of multimodal medical images (e.g., PET/CT, MRI). Its core capabilities include detection, spatial localisation, benign/malignant classification (including subtype differentiation), and prediction of key attributes (e.g., infiltration depth, molecular expression status) for target lesions (e.g., tumours, metastases). Within the specific context of radiopharmaceutical development, this module programmatically identifies metabolically active lesions in radiotracer images. It provides input for subtype classification models and supports efficacy prediction and recurrence risk assessment based on imaging features. Architecturally, this tool integrates advanced object recognition frameworks including Retina U-Net (employing a joint detection and segmentation architecture), the multi-task learning-suited nnDetection 53 framework, and CheXNet 28 (specialised for chest X-ray analysis). It also incorporates Transformer-based vision models (such as Vision Transformer 54 and Swin Transformer) to enhance performance in tumour classification and complex attribute prediction tasks. Methodologically, this module operates programmatically following tracer PET/CT examinations to perform lesion detection and localisation, calculate standard uptake values (SUV) and metabolic tissue volume (MTV) 55. These outputs serve as direct quantitative inputs for subsequent dosimetric calculations and treatment decision-making. To validate reliability, this study employs metrics including sensitivity, specificity, localisation error (mm), and classification accuracy.

Image Segmentation Module: This module is dedicated to the precise segmentation of target anatomical structures, lesions and their subregions from 2D/3D multimodal imaging data, providing the spatial foundation for subsequent quantitative analysis. It can process images with varying slice thicknesses and scanning protocols, generating high-quality masks to support accurate measurements of target volume, shape, position and other parameters. Within nuclear medicine research workflows, this module is primarily employed for lesion volume quantification, analysis of tracer distribution in target versus non-target regions, defining critical areas for dosimetry calculations, and preprocessing in vivo images. The core algorithm employs the adaptive nnU-Net framework, which programmatically optimises network architecture, preprocessing workflows, and training parameters for specific datasets. Its performance has been reported in multiple public challenges. Furthermore, this module integrates tools such as TotalSegmentator 56 (for multi-organ segmentation in whole-body CT/MRI) and Swin UNETR 57 (a Transformer-based 3D segmentation model) to address diverse segmentation requirements ranging from routine organs to complex lesions. In practical application, this tool generates contours for tumours and critical organs, reducing variability and time expenditure associated with manual delineation. Segmentation performance is quantified through metrics such as Dice Score and Hausdorff Distance 58, ensuring results are suitable for subsequent rigorous dosimetric and biodistribution analyses.

Image Enhancement Module: Addressing common issues in raw medical images such as low contrast, significant noise, and motion artefacts, this module is specifically designed to enhance image quality, thereby improving visual interpretation and the reliability of subsequent quantitative analysis. It achieves this through image processing techniques including denoising, contrast enhancement, and artefact correction, aiming to improve the visibility of minute lesion details. In nuclear medicine imaging scenarios, this is crucial for improving the signal-to-noise ratio of PET/SPECT images, increasing the detection rate of minute metastatic lesions, and enhancing the accuracy of pre- and post-treatment image comparisons. The module’s implementation primarily relies on Generative Adversarial Networks (GANs ^59^) and their variants, methodologies that have demonstrated significant potential in medical image enhancement, super-resolution, and artefact removal tasks. Concurrently, this tool integrates the enhancement algorithms into standardised medical image processing frameworks such as MONAI ^60^, ensuring compatibility and reproducibility of the workflow. Operationally, users may preprocess raw PET/CT or MR images prior to lesion identification or segmentation. Images enhanced through this module provide higher-quality input for downstream identification and segmentation tasks, thereby elevating the overall analytical workflow’s precision. Performance improvements can be demonstrated by comparing metrics such as the detection rate of minute lesions before and after enhancement.

Image Registration Module: To achieve spatial alignment of multimodal images (e.g., PET and CT) or multi-temporal images (e.g., baseline and follow-up scans), this module provides high-precision image registration capabilities. Its core function involves transforming images from different sources into a common coordinate system, enabling cross-modal quantitative comparisons and fusion analysis. Examples include overlaying tracer biodistribution maps onto detailed anatomical structures, or comparing pre-treatment dose distribution maps with post-treatment efficacy assessment images. This module integrates both deep learning-based registration networks (e.g., DFMIR 61, VMambaMorph 62) and traditional high-precision algorithm toolkits (e.g., ANTs 63, Elastix), supporting diverse transformation models ranging from rigid and affine to deformable registrations. In nuclear medicine research, this module supports precise matching between tracer kinetic images and high-resolution anatomical images, providing a spatially consistent foundation for target delineation and dose-volume histogram (DVH) analysis 64. Method descriptions must specify the registration strategy (e.g., rigid-then-deformable) and similarity measures employed. Registration accuracy is quantified through metrics such as the mean target registration error (TRE) between target point sets or mean surface distance, typically requiring sub-millimetre precision to meet the stringent demands of radiotherapy planning and quantitative analysis.

Pathological Image Diagnosis Module: To bridge microscopic histopathology with macroscopic molecular imaging, this module introduces weakly supervised deep learning analysis capabilities for whole slide images (WSI). This module enables automatic tissue region identification, lesion detection, tumour grading, and molecular subtype prediction without pixel-level annotation. This capability is crucial for verifying the target-binding specificity of radiopharmaceutical ligands and analysing the expression heterogeneity of target proteins within the tumour microenvironment during radiopharmaceutical development. At its core, this module integrates advanced models including CLAM 65 (a multi-instance learning framework based on Clustering-constrained Attention Mechanism) and CHIEF 66(a framework combining hierarchical feature extraction with context-aware processing). These models programmatically localise diagnostically relevant regions within whole-slide images and aggregate regional features for slide-level classification and prediction. In workflow integration, this module analyses immunohistochemistry (IHC) sections from the same patient to generate spatial distribution heatmaps of target expression. These are then spatially correlated with tracer uptake patterns from PET imaging, thereby validating radiological findings at the histological level. The efficacy of this approach is evaluated through metrics such as the area under the receiver operating characteristic curve (AUC) and F1 score, ensuring the reliability of pathological analysis and providing cross-scale biological interpretation for the mechanism of action of radiopharmaceuticals.

### Radiological Safety and Precision Dose Calculation Tools

RadGuide AI’s Radioactive Safety and Precise Dose Calculation Tool establishes a structured system for modelling the in vivo processes of radiopharmaceuticals and assessing their doses. By integrating standardised dose calculation software such as OLINDA/EXM and MIRDcalc with Monte Carlo simulation platforms including OpenTOPAS and GATE, it provides end-to-end support from radiopharmaceutical pharmacokinetic analysis to precise calculation of radiation doses to target tissues.Based on the Medical Internal Radiation Dose (MIRD) model, this tool precisely simulates the absorption, distribution, metabolism, and excretion of radionuclides within the body by inputting multi-modal imaging data such as PET/CT and SPECT. It quantitatively calculates radiation absorbed doses in both target and non-target tissues. In radiopharmaceutical development applications, this tool optimises radionuclide selection and labelling strategies, precisely evaluates target-to-non-target ratios (TNR), and validates clinical dose safety margins through voxel-level dose calculations.

Technically, the system employs OLINDA/EXM for standardised organ dose calculations, MIRDcalc for patient-specific absorbed-dose estimation, and OpenTOPAS or GATE for Monte Carlo simulation of particle transport and energy deposition. Outputs include dose-volume histograms, voxel-level dose maps, target/non-target absorbed-dose summaries, and safety-threshold alerts. The present manuscript treats these results as technical and preclinical feasibility evidence. Any patient-level dosimetric accuracy claim requires a prespecified cohort, measurement protocol, ground-truth definition, ethics approval, R2 with confidence intervals, MAE/NMAE, signed bias, and external validation.

The tool integrates CareSelect® Imaging’s examination suitability recommendation mechanism with the specialised logic of the NucMed clinical decision support system. Based on specific patient signs, imaging findings, and laboratory indicators, it provides decision-support suggestions for potentially appropriate nuclear medicine examination protocols and management options. Concurrently, the system incorporates the MDCalc risk assessment model, providing safety control functions such as drug interaction alerts and allergy risk notifications. Through process standardisation, it supports workflow standardisation and quality review. This integrated tool delivers comprehensive technical safeguards for the safety evaluation and individualized treatment evaluation of radiopharmaceuticals, supporting standardised and individualised nuclear medicine evaluation.

### Performance Evaluation of RadGuide AI

As illustrated in Figure 7, RadGuide AI’s intelligent agent workflow demonstrates a traceable interaction process from clinical query input to structured decision output. The process commences with natural language queries and integrated analysis. When clinicians describe clinical requirements using everyday diagnostic language, the system first employs natural language understanding technology to accurately interpret query intent. It then utilises a medical terminology standardisation module to ensure precise comprehension of specialised concepts (e.g., “hepato-pulmonary shunt ratio”, “target-to-background ratio”). During the intelligent retrieval and evidence integration phase, the system employs the RAGFlow tool to execute multi-level knowledge retrieval. Through the hierarchical perceptual retrieval strategy of the medical expert system, it acquires structured evidence from diverse sources including clinical guidelines, case reports, and patent literature. Medical entity recognition then structurally encapsulates the retrieval results, supporting the authority and relevance of the information.

**Figure 7.**
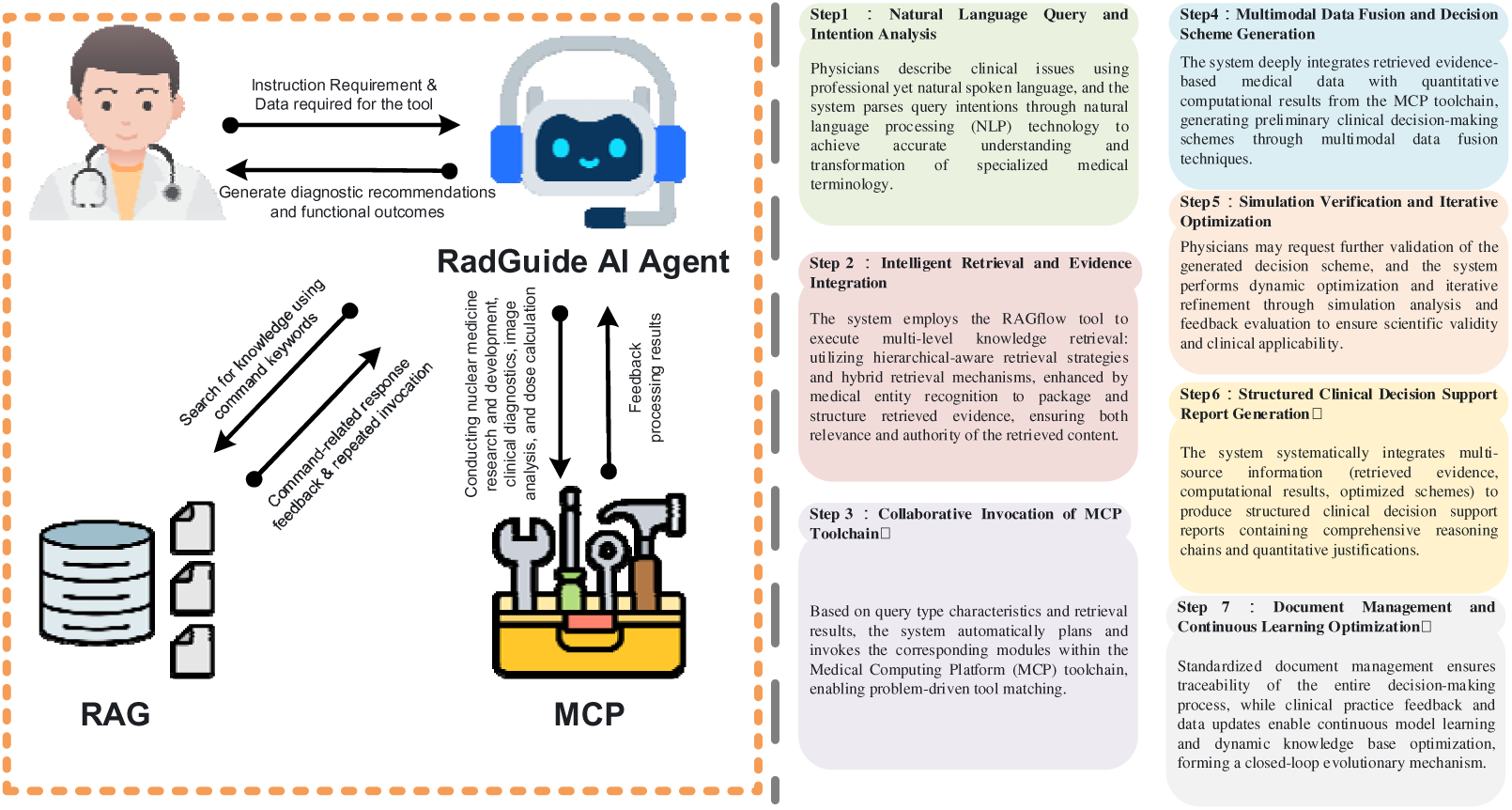
Detailed usage workflow of RadGuide AI

Based on query characteristics and retrieval outcomes, the system enters the collaborative invocation of the MCP toolchain phase, planning and invoking corresponding modules within the medical computing platform. For instance, when evaluating ¹LLLu-PSMA treatment protocols, the system may concurrently invoke tools such as the “Organ Dose Calculation Tool,” “Pharmacokinetic Simulation Tool,” and “Image Registration Tool,” achieving problem-driven intelligent tool matching. Subsequently, during the multimodal data fusion and decision-making scheme generation phase, the system integrates retrieved evidence-based medical data with tool computation results. Through cross-modal information alignment technology, it generates preliminary decision-support summaries, such as draft treatment-support recommendations for qualified review incorporating specific administration doses, imaging time windows, and safety thresholds.

To support workflow applicability while maintaining safety boundaries, users may request further verification of generated plans through Monte Carlo simulation, MIRD-based cross-checking, and rule-based threshold checks. The system then produces a structured decision-support report containing retrieved evidence, computation results, uncertainty notes, and the audit trail. Recommendations are not intended for autonomous clinical use.

Finally, within the documentation management and continuous learning optimisation phase, standardised document management supports traceability throughout the decision-making process. Clinical practice feedback and data updates drive the model’s continuous learning and dynamic knowledge base refinement, establishing a closed-loop evolutionary mechanism: “clinical input – intelligent processing – practice feedback”. This process design not only achieves traceable conversion from natural-language inputs to decision-support summaries but also transforms the traditional, expert-dependent nuclear medicine decision-making process into a standardised, reproducible intelligent workflow through three key mechanisms: tool collaboration, multimodal fusion, and iterative optimisation. This supports evaluation of workflow efficiency and reliability under human review.

#### MCP Tool Use Case Analysis

This section presents case studies of the MCP tool, encompassing two domains: radiopharmaceutical development and radiation safety with precise dosing.

#### Case Study 1: Radiopharmaceutical Development – An Example of Radiotargeted Drug Development

Radioactive pharmaceuticals play a vital role in tumour diagnosis and targeted therapy. However, their development involves lengthy cycles, high experimental costs, and reliance on experience and repetitive experiments during target screening, drug conjugation, and radionuclide selection. Key challenges include: difficulty in target identification due to complex radioligand binding sites arising from pathological heterogeneity; uncertainty in drug conjugation efficiency and stability, with varying linkers affecting affinity and metabolic half-life; and safety risks in radionuclide selection and dose control, necessitating a balance between imaging signal and radiation safety.

Based on the RadGuide AI MCP toolkit, this study established a structured research and development workflow for radiopharmaceuticals, as illustrated in Figure 8. The system commences with TCGA 67 pan-cancer transcriptomics data, employing differential expression analysis and LASSO regression 68 to identify 110 core tumour biomarkers. These are cross-validated against the Labome Antibody Database, ultimately screening 66 candidate target genes supported by commercially available antibodies to ensure the clinical translational potential of target selection. During the molecular design phase, the system integrates multi-platform tools including AutoDock Vina, RosettaLigand, and MOE for ligand-protein binding simulations. Combining quantitative structure-activity relationships (QSAR), ADMET property optimisation, it generates a molecular conformation database incorporating linkers such as PEG and DOTA, providing candidate chemical design options for subsequent radionuclide labelling. The pathological imaging validation module uses the CLAM/CHIEF weakly supervised learning framework for automated tissue segmentation and quantitative analysis of target expression in immunohistochemical sections. Concurrently, Retina U-Net and Swin Transformer models enable automatic lesion detection and target/non-target uptake ratio calculation within PET/CT images, supporting a workflow from molecular design to in vivo validation. This workflow employs an intelligent engine to programmatically coordinate three major processes: target screening, molecular design, and imaging validation. It enhances the standardisation and iterative efficiency of radiopharmaceutical development, providing a reproducible, computation-driven R&D paradigm for nuclear medicine research.

**Figure 8.**
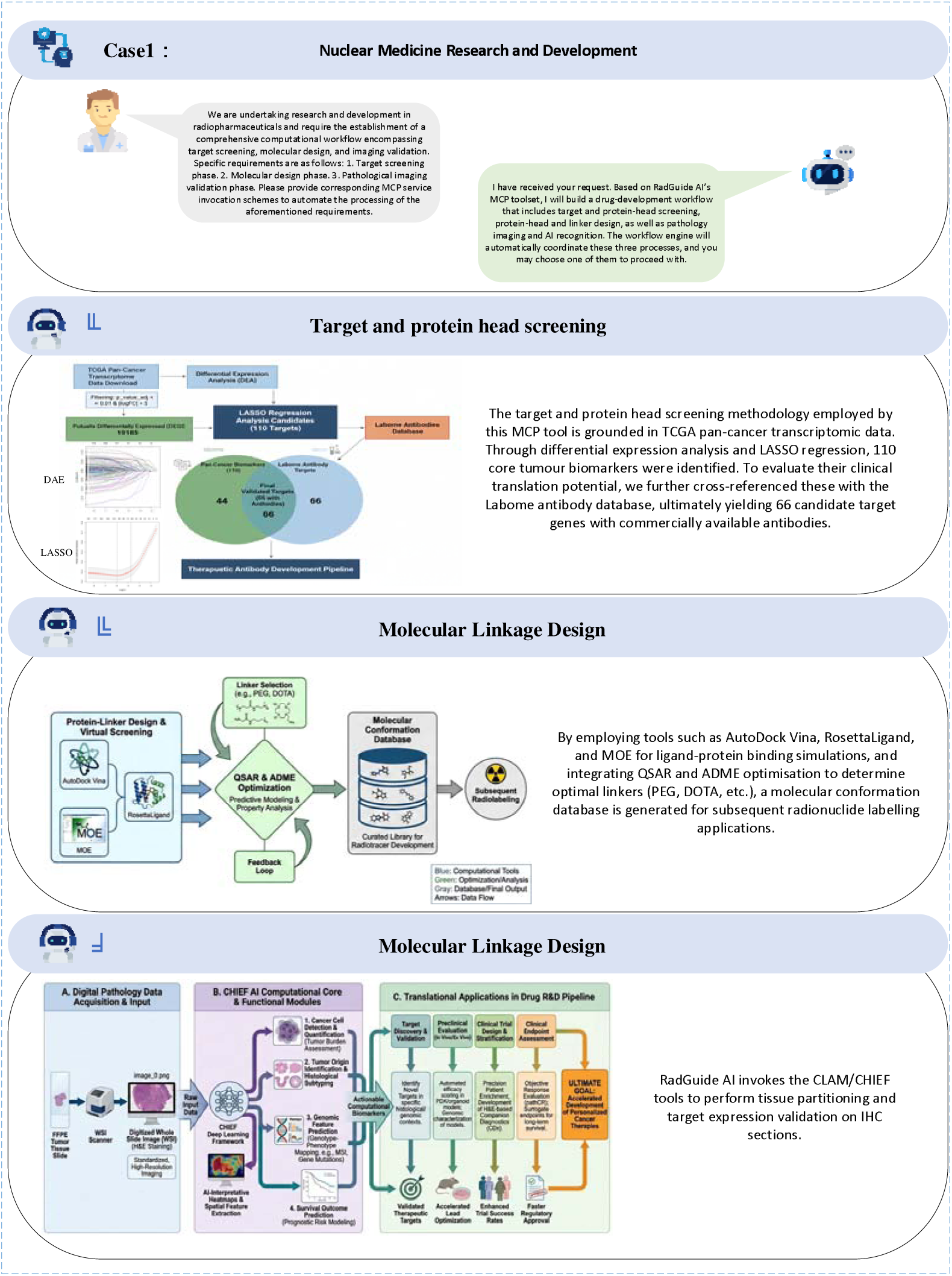
Nuclear medicine development case studies

### Case Study Two: Radiological Safety and Precision Dosing

#### Patient-Specific Radiopharmaceutical Dose Optimisation

Radiopharmaceuticals are extensively utilised in tumour diagnosis and treatment ^69–71^, yet dose design faces two primary challenges:

1. **Safety concerns**: Complex distribution of radionuclides within the body may result in high doses being absorbed by non-target organs, increasing the risk of toxic side effects;
2. **Insufficient Dose Precision**: Conventional dose calculations rely on standardised models, struggling to account for individual variations (body weight, organ size, metabolic rate, etc.), thereby compromising therapeutic efficacy and safety.

To achieve safe and effective personalised radiopharmaceutical therapy, we propose the MCP tool for radiological safety and precision dose calculation, integrating image processing, dose simulation, and clinical decision-making.

Figure 9 illustrates RadGuide AI’s radiation safety and precision dose calculation case study, presenting the traceable computational workflow for individualised dose optimisation of ¹LLLu-labelled therapeutics in mice. This process commences with interaction between the intelligent agent and the user. Upon receiving a dose calculation request, the system programmatically identifies and prompts for critical parameters such as mouse body weight and tumour volume, demonstrating the tool’s intelligent validation capability for input completeness. At the core of dose calculation, the tool employs a three-tier progressive computation strategy: First, it calculates the theoretical activity of the target region based on the MIRD model. Using the formula AMIRD = CFto177, tumor Dtumor × Mtumor, it determines the minimum ¹LLLu activity (≥0·03 MBq) required to achieve an 80 Gy tumour dose. Secondly, an organ uptake scoring system is established. Relative organ uptake is quantified using the formula forgan = ∑i Countsi Countsorgan. This is combined with organ mass and dose conversion factors to calculate maximum permissible activities for critical organs such as the kidneys and liver.Ultimately, the physiologically acceptable activity range is determined via the composite constraint function Aphys = min(AMIRD, Amax, kidney, Amax, liver, Abot). Following safety factor adjustment, the final recommended injection activity is output (2·9–3·0 MBq).

**Figure 9.**
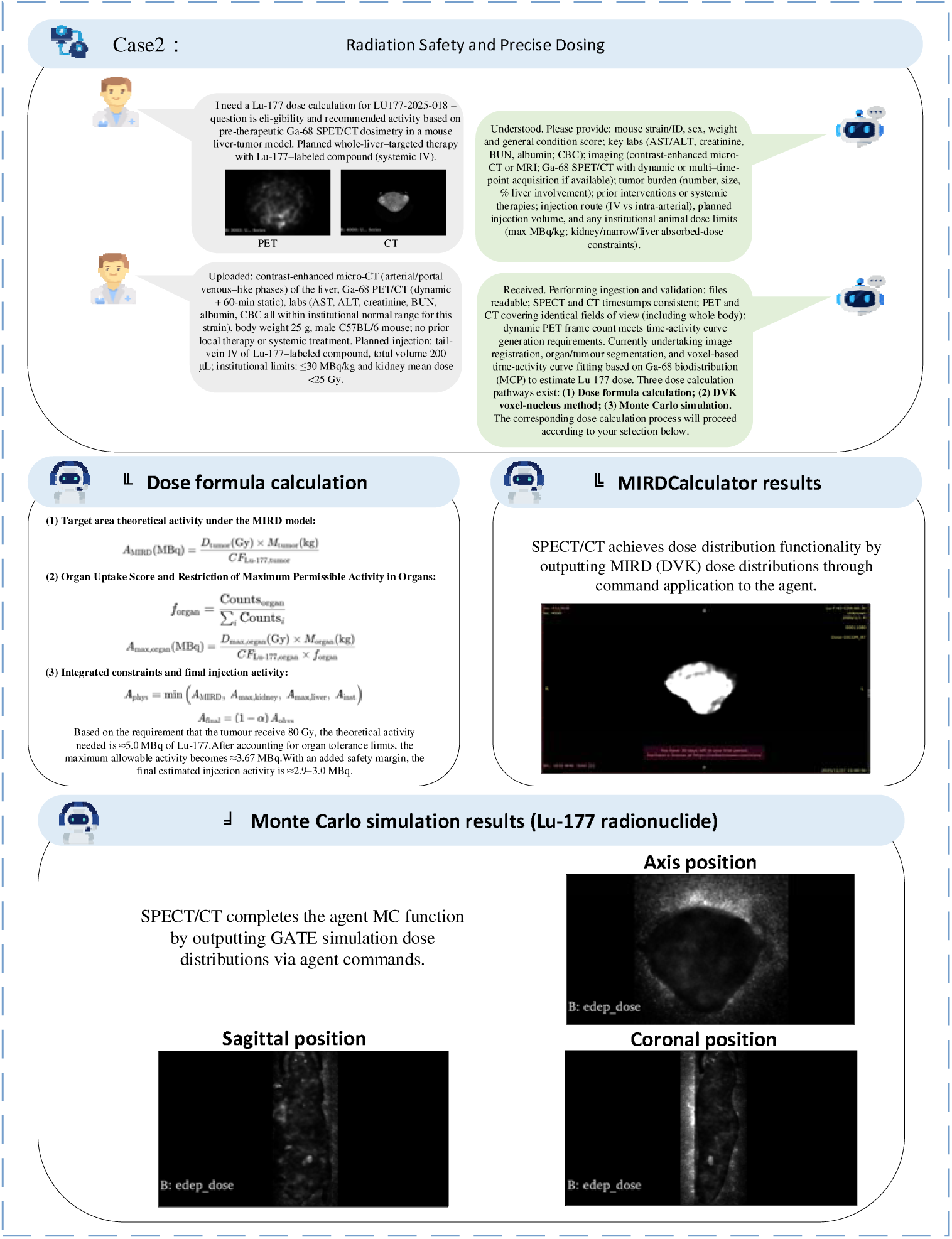
Radiological safety and precision dose-calculation case study

This case study demonstrates the dual validation mechanism of MIRDCalculator and Monte Carlo simulation. The MIRDCalculator module processes SPECT/CT image data to generate dose-volume histograms (DVHs) and three-dimensional dose distributions, supporting dose assessment based on standard models. The Monte Carlo simulation module, integrated with the GATE platform, performs physics-based simulations of the transport process of ¹LLLu decay particles within the anatomical structures of real mice. It outputs voxel-level dose distribution maps in the sagittal, axial, and coronal planes, visually illustrating the deposition differences of the radionuclide in tumour and normal tissues. This multi-methodological cross-validation design supports computational efficiency while supporting the physical accuracy of dose predictions through first-principles simulations, providing a reliable technical pathway for dose optimisation in preclinical research.

RadGuide AI’s radiation safety and precision dose calculation tool achieves personalised radiopharmaceutical dose optimisation by establishing a structured workflow encompassing multimodal data, activity distribution, dose simulation, and clinical decision-making. This Medical Computing Protocol MCP tool offers systematic solutions to two core challenges in radiopharmaceutical dose design: non-target organ safety risks and inadequate adaptation to individual variability. The tool first integrates multi-modal imaging data (PET/CT, SPECT) and patient clinical parameters. Using MCP medical imaging processing tools, it performs automated pre-processing, segmentation, and multi-modal registration to establish a standardised voxel-level data input format. During activity distribution and lesion identification, the system employs deep learning models such as Retina U-Net and Swin Transformer to programmatically detect lesions and quantify tracer uptake. This is combined with the CLAM/CHIEF pathology recognition module for cross-scale target expression validation, ensuring consistency between biological targeting and imaging manifestations.

Dose simulation and optimisation employ OpenTOPAS and GATE to model radionuclide energy deposition in individualised or preclinical phantoms. MIRDcalc/OLINDA/EXM supports organ absorbed-dose calculation and multi-objective optimisation. The output includes voxel dose maps, target/non-target absorbed-dose ratios, organ dose limits, and recommended protocol ranges. Patient-level R2 claims are not made unless cohort size, measurement protocol, and ethics documentation are provided.

### Technical Validation and Feasibility Evaluation

RadGuide AI was evaluated as a technical and feasibility-stage system rather than as a prospectively validated clinical device. The validation framework includes benchmark-based model evaluation, tool-routing and execution auditing, factuality and numerical-error checks, and preclinical workflow demonstrations. Guideline-related evaluation uses MIRD dosimetry principles, EANM guidance, and relevant SNMMI/ACR materials as reference sources where applicable.

For radiopharmaceutical development, RadGuide AI was tested in workflow demonstrations covering target screening, molecular design, linker evaluation, radionuclide selection, image-analysis support, and preclinical dose optimisation. For dosimetry, the system was evaluated through demonstrative 177Lu workflows using MIRD-based calculations and Monte Carlo cross-checking in mice or phantom-like settings, as shown in Figure 9. No completed human clinical validation at Zhongda Hospital is claimed in the present report; such claims require the corresponding cohort, measurement protocol, ground truth, statistical analysis, and ethics approval.

The current evidence supports feasibility of the data-model-tool architecture and identifies the reporting requirements for future validation. Future patient-level studies should prospectively define cohort size, inclusion/exclusion criteria, treatment setting, imaging schedule, dosimetry ground truth, safety endpoints, inter-rater or inter-operator variability, and ethics/IRB approval. Until such studies are completed, RadGuide AI should be described as a research and decision-support prototype.

## Conclusion

RadGuide AI demonstrates that a nuclear medicine agent can integrate multi-source curated data, a domain-adapted large language model, and specialised toolchains to provide transparent and auditable support for nuclear medicine R&D workflows. The curated core dataset comprises 5,474 high-quality QA items after progressive quality control and semantic deduplication. In the locked N=200 technical evaluation set, RadGuide-LLM achieved an answer accuracy of 88.5% and a Macro-Average score of 21.5/25. These findings support feasibility of domain adaptation while further independent validation is required.

The 55 specialised nuclear medicine tools integrated via MCP support transformation from natural-language queries to executable workflows, including target screening, molecular design, image analysis, dose calculation, and safety assessment. The case studies demonstrate feasibility for radiopharmaceutical development and preclinical dose-optimisation workflows. Quantitative claims about clinical effectiveness, patient-level dose accuracy, and workflow time savings have been restricted to settings where denominators, ground truth, and statistical procedures are specified.

The current study remains limited by its feasibility-stage design, reliance on generated and curated benchmark data, and absence of a prospective multicentre patient-level validation cohort. Future work will focus on independent human-curated benchmarks, formal ablation experiments, external validation of imaging and pathology modules, prospective dosimetry studies with ethics approval, uncertainty quantification, and public release of reproducibility artefacts where licensing and privacy constraints permit.

## Discussion

This study developed RadGuide AI, a nuclear medicine agent intended to connect curated domain data, language-model reasoning, and specialised computational tools through a traceable data-model-tool framework. The core contribution is an implementation pathway for auditable decision support in which natural-language intent can be translated into evidence retrieval, tool invocation, verification, and structured reporting. The evidence base is positioned as technical evaluation rather than completed clinical validation.

Several limitations remain. First, the benchmark includes LLM-generated QA items and requires expansion with independent human-curated cases. Second, the current toolchain evaluation is feasibility-oriented; formal measurements of tool-routing accuracy, execution success, operator workload, and failure modes should be reported in future studies. Third, off-the-shelf imaging and pathology models may not generalise across scanners, staining protocols, disease types, or institutions without external validation. Fourth, hallucination risk remains clinically important; future evaluations should quantify fabricated citations, unsupported numerical parameters, unit errors, and citation-faithfulness failures. Human-in-the-loop review is mandatory for safety-critical use.

Looking ahead, RadGuide AI’s technical trajectory holds potential for broader application. Firstly, its core architecture is adaptable to other radionuclide therapy scenarios, such as ²²LAc-targeted therapy or LLCu-diagnostic imaging. Secondly, its integrated toolkit model provides a technical framework for intelligent assessment of combined drug-device therapies. Crucially, the system’s established “benchmark-driven optimisation” methodology offers a reference for developing AI systems in other medical specialties. Key priorities for subsequent work include conducting multicentre prospective clinical validation, deepening research into uncertainty quantification mechanisms, and establishing a dynamic knowledge base update system to further enhance the system’s research and decision-support utility and timeliness.

In summary, RadGuide AI represents a feasible framework for traceable intelligent nuclear medicine research support. By integrating expert knowledge, computational tools, and workflow audit logs, it offers a path from concept generation to structured technical evaluation. Broad clinical deployment will require prospective multicentre validation, transparent benchmark release, external tool-module validation, and governance mechanisms for safety, privacy, and accountability.

## Declaration of interests

We declare no competing interests.

## Data availability statement

The data supporting the findings and technical evaluation of this study are available from the corresponding author upon reasonable request, subject to licensing, privacy, and institutional governance restrictions.

## Consent for publication

All participants signed informed consent.

## Acknowledgments

This work was supported by the National Natural Science Foundation of China (82130060), the Jiangsu Provincial Basic Research Program Natural Science Foundation–Frontier Leading Technology Basic Research Project (BK20232008), the Jiangsu Provincial Medical Innovation Center (CXZX202219), and the Jiangsu Provincial Major Science and Technology Project (BG2024007). Institutional and clinical research support was provided by Zhongda Hospital, Medical School, Southeast University.

We thank the clinicians, medical physicists, radiologists, and researchers from the Center of Interventional Radiology and Vascular Surgery and collaborating departments at Zhongda Hospital who contributed technical discussions, workflow feedback, and domain expertise. Any future use of patient-level data for clinical validation will require separate ethics/IRB approval and will be reported with cohort, protocol, and statistical details.

We additionally acknowledge the developers and maintainers of the open-source software packages and computational tools used and cited in this publication, whose contributions were essential to data processing, modelling, and analysis. Zhongda Hospital and Southeast University are supported by provincial and national research and education funding programmes.

## Notes

The study was supported by the National Natural Science Foundation of China (82130060), Jiangsu Provincial Basic Research Program Natural Science Foundation-Frontier Leading Technology Basic Research Project (BK20232008), Jiangsu Provincial Medical Innovation Center (CXZX202219), and the Natural Science Foundation of Jiangsu Province (BG2024007). The funding sources had no role in the writing of the report or in the decision to submit the paper for publication.

### Competing Interest Statement

The authors have declared no competing interest.

